# GLIOBLASTOMA MULTIFORME: A META-ANALYSIS OF DRIVER GENES, CURRENT DIAGNOSIS, AND TUMOR HETEROGENEITY

**DOI:** 10.1101/2020.10.19.20215467

**Authors:** Gabriel Emilio Herrera-Oropeza, Carla Angulo-Rojo, Santos Alberto Gástelum-López, Alfredo Varela-Echavarría, Maribel Hernández-Rosales, Katia Aviña-Padilla

## Abstract

Glioblastoma (GBM) is the most aggressive and common brain cancer in adults with the lowest life expectancy. The current neuro-oncology practice has incorporated genes involved in key molecular events that drive GBM tumorigenesis as biomarkers to guide diagnosis and design treatment. This study summarizes findings describing the significant heterogeneity of GBM at the transcriptional and genomic levels, emphasizing eighteen driver genes with clinical relevance. A pattern was identified fitting the stem cell model for GBM ontogenesis, with an up-regulation profile for *MGMT* and down-regulation for *ATRX, H3F3A, TP53*, and *EGFR* in the mesenchymal subtype. We also detected overexpression of *EGFR, NES, VIM*, and *TP53* in the classical subtype and of *MKi67* and *OLIG2* genes in the proneural subtype. In keeping with this, we found a panel of nine biomarkers with a strong potential to determine the GBM molecular subtype. A unique distribution of somatic mutations was found for the young and adult population, particularly for genes related to DNA repair and chromatin remodeling, highlighting *ATRX, MGMT*, and *IDH1*. Our results also revealed that highly lesioned genes undergo differential regulation with particular biological pathways for young patients. This meta-analysis will help delineate future strategies related to the use of these molecular markers for clinical decision-making in the medical routine.

## Introduction

Glioblastoma multiforme (GBM) is the most frequent and aggressive deadly primary brain tumor in adults, accounting for approximately 82% of all malignant gliomas(1). Although it can affect children, its incidence rises with age. GBM tumors are characterized by increased cell proliferation, aggressive invasion, active angiogenesis, and a remarkable genetic heterogeneity (2). Histologically, tumors display a high morphological variability as they contain pleomorphic and multinucleated cells with high mitotic activity, show microvascular proliferation, undergo severe and characteristic endothelial hyperplasia, contain intravascular microthrombi and extensive necrosis of an ischemic or pseudo-empalized nature. The multiforme denomination of GBM tumors is due to the diverse and heterogeneous microenvironments that parallel their multiple histological patterns and cytological features.

According to their ontogeny, most GBMs are primary tumors that develop de novo in the absence of previous neoplasia. Primary GBMs are highly aggressive and invasive, tend to extend to both cerebral hemispheres, or are bilateral, and they are most commonly manifested in elderly patients. Secondary GBMs, in contrast, are located in the frontal lobe and develop mainly in younger patients suffering from anaplastic astrocytoma or low-grade astrocytoma, presenting a much better prognosis (3). Recent reports have determined that primary and secondary glioblastomas have distinct genetic alterations related to particular biological pathways (1, 3, 4), suggesting they require different therapeutic approaches. Hence, from the clinical perspective, discerning between primary and secondary GBM is highly relevant (2). Usually, primary GBMs present overexpression and gene amplification of epidermal growth factor receptor (*EGFR*) and mutations in cyclin-dependent kinase inhibitor 2A (*CDKN2A/p16INK4A*) and phosphatase tensin homolog (*PTEN*) genes. Molecular biomarkers of secondary GBM include mutations in tumor protein 53 (*TP53*) and isocitrate dehydrogenase-1 (*IDH1*) genes, which correlate strongly with O6-Methylguanine-DNA methyltransferase (*MGMT*) promoter methylation (3, 5).

Initiation and progression of GBM tumorigenesis are related to genetic and epigenetic alterations and molecular subtypes of GBM have unique transcriptional profiles. Based on expression features, GBM tumors were originally classified into four subtypes: Neural, Proneural, Classical (proliferative), and Mesenchymal (6), a scheme that has been recently revised using transcriptomic information. The improved classification eliminates the neural subtype and considers tumors of this molecular type as containing normal brain tissue contamination (7, 8). GBM molecular subtypes are also associated with different spatial zones, heterogeneity, and aggressiveness of the tumor (9).

GBMs belonging to the proneural subtype have alterations in *TP53, PDGFRA, PIK3CA*, and *IDH1* genes (10, 12). The classical subtype, also known as a proliferative subtype, has been associated with high levels of cell proliferation and upregulation of *EGFR*(11). Mesenchymal GBMs show overexpression of mesenchymal and astrocytic markers (*CD44*, and *MERTK*) and down-regulation of neurofibromatosis type-1 (NF1) and up-regulation of chitinase 3 like 1 (*CHI3L1/YKL-40*) and *MET* genes are frequently observed (10). While the proneural subtype has been mostly reported in younger patients and is associated with a favorable prognosis, the mesenchymal and the classical subtypes are usually linked to more aggressive high-grade gliomas that appear in adult or elderly life.

Recent advances employing Next Generation Sequencing have led to a better insight into the molecular biology of gliomas contributing potential markers for better diagnosis, and new approaches to finding specific treatment strategies (13). GBM remains an incurable deadly disease with an abysmal prognosis that has not significantly shown improvement, causing an enormous individual and societal burden. Thus, there is a need for tumor-specific drug targets and pharmacological agents to inhibit cell migration, dispersal, and angiogenesis (7).

GBM heterogeneity makes this type of cancer one of the most challenging to treat and consists of *inter-tumor* and *intra-tumor* feature variations. Inter-tumor heterogeneity refers to GBMs from different patients with altered and differing genotypes and phenotypes related to diverse etiological and environmental factors. On the other hand, intra-tumor heterogeneity refers to the presence of multiple and different cell subpopulations within the same tumor, defining its topology and architecture (14). The comprehensive genomic classification of GBM paves the way for an improved understanding of GBM, which in the future may result in personalized therapy. Hence, there is an urgent need to further our knowledge of tumor heterogeneity as it will help design better therapies against GBM and tumor recurrence.

Based on a meta-analysis, in this study, we describe the heterogeneity of GBM at the transcriptional and genomic levels, with emphasis on driver genes currently used as genomic markers. For that purpose, from sixty clinical reports, we selected and analyzed eighteen driver genes that have shown deregulated behavior in patient samples. Using bioinformatics pipelines, we examined their mRNA expression in the different GBM molecular subtypes and the presence of somatic mutations linked to possible disruption of protein function. We hope that the new knowledge generated in this study leads to novel therapeutic intervention strategies.

## Materials and methods

### Data mining and selection of GBM driver genes currently used as genomic markers in the clinic

Literature research was performed using a systematic approach to identify GBM biomarkers in the routine clinical diagnosis that yielded differential transcriptomics or genomic profiles on tumor samples. Using a combination of three terms (1) *“Glioblastoma”*, (2) *“Clinical”, and* (3) “*Case”*, a total of 3,238 clinical reports were found using the BVS (1548), Cochrane (0), Karger (271), and PubMed (1419) databases. Clinical reports were identified and selected by title and summary. All articles were evaluated using the guidelines of the Preferred Reporting Items for Systematic Reviews and Meta-Analyses (PRISMA) (http://prisma-statement.org/) to determine their eligibility, resulting in sixty reports, as described in Suppl. Material File 1. The search was conducted in July 2020 and focused on studies published in June 2000 – June 2020.

### Data source for the gene expression analysis

Eighteen genes were found to be involved in GBM diagnosis during the neuro-oncology clinical routine and evaluated for their mRNA expression analysis using data from the Glioblastoma BioDiscovery Portal (GBM-BioDP) https://gbm-biodp.nci.nih.gov (15). The gene expression data includes normalized (level 3) data from Verhaak 840 Core, a filtered data set conformed of three microarray platforms: HT_HG-U133A (488 patient samples/ 612042 features), HuEx-1_0-st-v2 (437 patient samples /618631), and AgilentG4502A_07_1/2 (101+396 patient samples /617813). GBM molecular subtypes were assigned according to the Verhaak classification (12).

### Determination of gene expression of GBM driver genes

We classified the mRNA expression analysis of the driver genes according to their biological ontology into three groups: 1) DNA repair and chromatin remodeling, 2) Cytoskeleton and cellular proliferation, and 3) Tumor suppressors genes. Using Python scripts (https://github.com/kap8416/GBM-META-ANALYSIS-OF-DRIVER-GENES), we determined the average and standard deviation of the z-score expression values for all patient results and classified them into the molecular GBM subtypes (Classic, Proneural, Mesenchymal, and Neural), and grouped them by their corresponding biological gene ontology group. Although the Neural subtype was eliminated in the improved classification, we included it in this study and considered it as normal tissue.

We examined the mRNA expression patterns of the driver genes clustered by patient subgroups taking their age into account. Three subgroups were created: 10-29-year-old patients (young subgroup), 30-59-years-old (adult subgroup), and 60-89-years-old (elderly subgroup). The average of the z-score values among the patient subgroups was clustered into the molecular GBM subtypes.

Finally, the Mann-Whitney test was used to examine the statistical difference in the mRNA expression z-scores between GBM molecular subtypes and patient subgroups and between each gene and GBM subtypes. Statistical significance for the test was set to *p < 0*.*05*.

### Data source for somatic mutations of GBM

Genomic data for the eighteen genes previously identified as molecular markers was downloaded from the NIH website https://portal.gdc.cancer.gov/ using the following restriction criteria: Primary site: brain; Program: TCGA; Project: TCGA-GB; Disease Type: gliomas; Sample type: primary tumor; Clinical age of diagnosis: 10-29 years, 30-59 years, and 60-89 years.

### Determination of mutations in GBM and driver genes

Using Python scripts (https://github.com/kap8416/GBM-META-ANALYSIS-OF-DRIVER-GENES), the number of mutations per gene in the TCGA-GBM project was determined by calculating the amount of different genomic DNA changes reported in each gene. Subsequently, the relative percentage of mutations per chromosome was calculated by taking into account the length and the number of coding genes of the respective chromosome. Substitutions, deletions, and insertions were identified, then the number of nucleotide changes occurring in all genes was determined, and their distribution was compared to the distribution of those present in the driver genes. Moreover, the total number of mutations per gene and the genome location of the somatic mutations were compared among patient subgroups according to their age. Finally, the protein phenotype impact values (polyphen) of all the canonical missense variant consequences of the driver genes in the TCGA GBM project were determined, analyzed, and compared between patient subgroups clustered by age.

### Functional enrichment for driver genes, unique or shared pathways

GO enrichment analysis was performed using the Metascape tool (http://metascape.org/). We then used the meta-analysis workflow to compare the driver gene pathways with those of the highly mutated genes to identify unique or shared biological pathways in which they are involved. Using Python scripts the top fifty mutated genes observed in the TCGA-GBM project were clustered by age group. Those genes were selected and analyzed by their GO enriched terms. Finally, affected genes in their protein polymorphism phenotype with >3 probably damaging consequences (PR) were clustered by the patient subgroups for the GO terms and TRRUST enrichment analysis.

## Results

### Identification of GBM driver genes as genomic markers in the clinic

Sixty clinical reports were found from 2005 to 2020 that were fully-text reviewed. A total of 73 patients with GBM were characterized and described in Table 1. Patient demographics consisted of 43 men and 30 women with ages ranging from four to seventy-eight years and a mean age of 43.31. Twenty-two patients were classified as young (4-29 years), thirty as adults (30-59 years), and twenty-one as elderly (60-78 years).

**Table 1.**
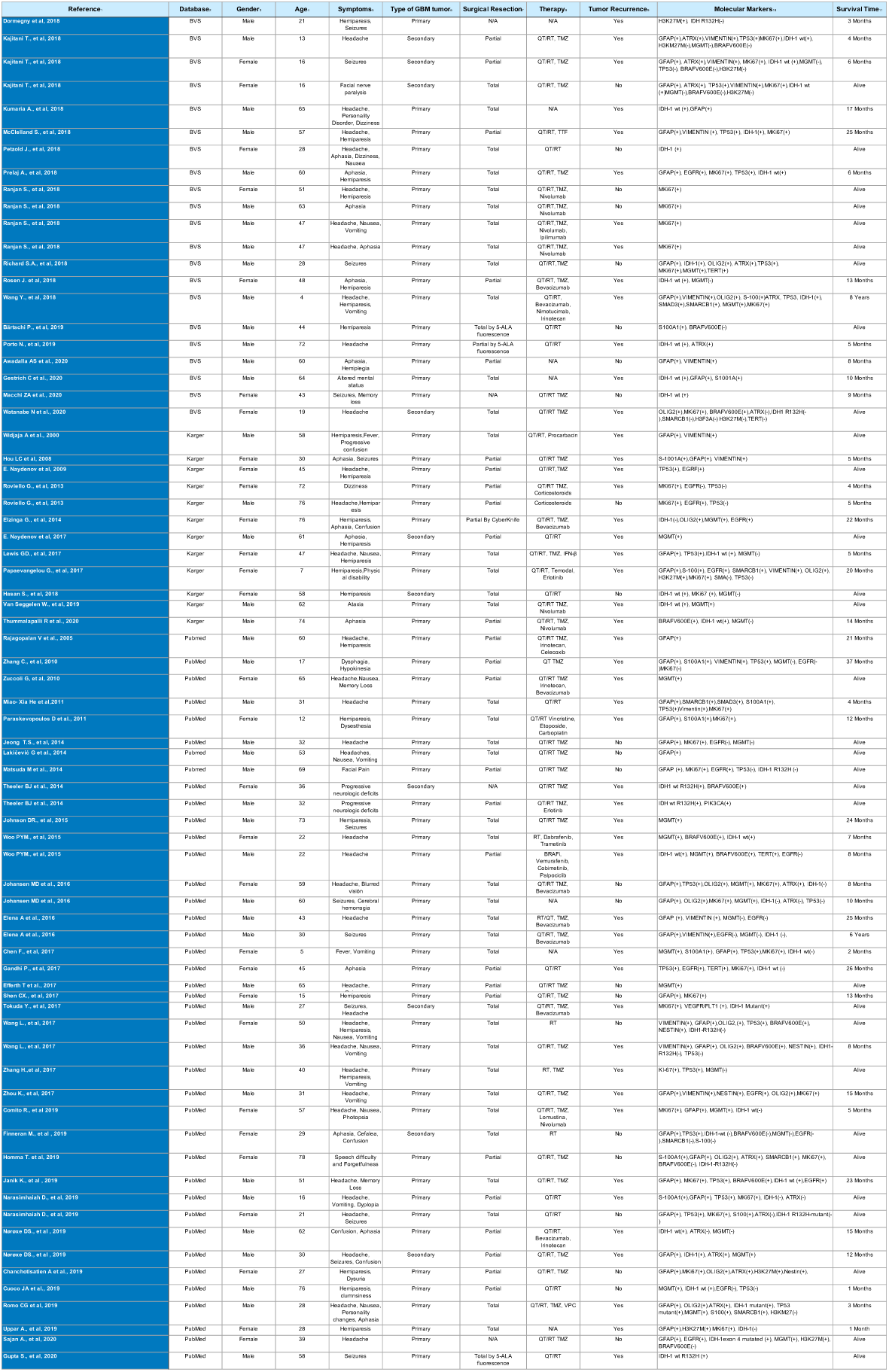
Summary of selected clinical cases of GBM.

Patients underwent a biopsy procedure to evaluate the expression and mutations of biomarkers, which were the most representative genes used in clinical cases over the last fifteen years. More than 80% of the clinical cases highlighted the use of a combination of two to eleven of the eighteen genomic markers. The most-reported were *IDH1, GFAP, MKi67*, and *MGMT*, followed by *TP53, ATRX*, and *EGFR*.

In this meta-analysis, only the biomarkers with differential positive results for patient diagnosis in the clinical reports were selected for further analysis (Table 1). According to their Biological Process Gene Ontology, driver genes were clustered using k-means into three groups to determine their possible role in common pathways. The first group includes *ATRX, H3F3A, IDH1, MGTM*, and *TERT* driver genes related to DNA repair and involved in chromatin remodeling pathways. The second group includes the cytoskeleton and cellular proliferation-related genes *EGFR, FLT1/(VEGFR), BRAF, GFAP, MKi67, NES, OLIG2, PIK3CA, SMAD3, S1001A*, and *VIM*. In particular, *EGFR* has an essential role in activating the receptor tyrosine *kinase/Ras/phosphoinositide3-kinase RTK/RAS/PI3K* pathway. Alterations in this pathway disrupt the G1-S transition in the cell cycle, which is highly relevant in the progression and excessive proliferation of GBM tumor cells. The third group included tumor suppressor genes *SMARCB1/INI1* and *TP53* which are negative regulators of cell growth control, normally acting to inhibit tumor development.

### Differential expression meta-analysis of driver genes of GBM tumorigenesis

Due to the high inter- and intratumor heterogeneity in GBM and to gain insight into this complex process, the expression profiling pattern of the top eighteen genes used as biomarkers in the clinical report meta-analysis was analyzed using gene expression data from the Glioblastoma BioDiscovery Portal. We focused on this analysis according to the Verhaak molecular classification of GBM, which groups tumors as neural, proneural, classical, and mesenchymal (12). Although an improved classification recommends no to include the neural subtype, we decided to use it as reference of non or low-proliferative tissue, also denominated as normal tissue as it derives from the PBZ (peripheral brain zone)(14). The gene expression analysis included all data available from the GBM-BioDP, including 370 samples for *NES* and *OLIG2* genes, 270 samples for *EGFR* gene, and 197 samples for the other driver genes (Table 2).

**Table 2.** Summary of driver gene expression in GBM molecular subtypes with significant *p*-value.

|  | NEURAL | PRONEURAL | CLASSICAL | MESENCHYMAL |
| --- | --- | --- | --- | --- |
| ***DNA Repair and Chromatin remodeling genes*** |  |  |  |  |
| ATRX | **-0.176** ± 0.761 | **0.37** ± 0.936 | **0.084** ± 0.601 | **-0.213** ± 0.591 |
| BRAF | **-0.378** ± 0.378 | **-0.152** ± 0.495 | **-0.237** ± 0.563 | **0.056** ± 0.468 |
| H3F3A | **-0.038** ± 0.548 | **0.34** ± 0.572 | **-0.079** ± 0.613 | **-0.495** ± 0.606 |
| MGMT | **0.716** ± 1.716 | **-0.128** ± 1.237 | **-0.078** ± 1.464 | **0.614** ± 1.268 |
| TERT | **0.099** ± 0.422 | **0.148** ± 0.385 | **0.26** ± 0.482 | **0.156** ± 0.49 |
| ***Cytoskeleton and celular proliferation genes*** |  |  |  |  |
| EGFR | **0.865** ± 3.77 | **-3.494** ± 3.78 | **3.502** ± 4.36 | **-2.002** ± 3.787 |
| FLT1 | **-0.824** ± 1.06 | **-0.571** ± 0.813 | **-0.301** ± 1.023 | **0.082** ± 1.093 |
| GFAP | **0.233** ± 0.664 | **0.114** ± 0.87 | **0.367** ± 0.493 | **-0.293** ± 1.037 |
| IDH1 | **0.224** ± 0.715 | **-0.175** ± 0.881 | **0.484** ± 1.089 | **-0.168** ± 0.872 |
| MKI67 | **-1.072** ± 1.292 | **1.019** ± 1.545 | **-0.114** ± 1.269 | **-0.325** ± 1.005 |
| NES | **-0.745** ± 1.025 | **-0.032** ± 0.852 | **1.525** ± 1.004 | **0.053** ± 0.909 |
| OLIG2 | **-0.052** ± 1.051 | **1.316** ± 1.182 | **0.07** ± 1.173 | **-1.455** ± 0.964 |
| PIK3CA | **-0.92** ± 0.595 | **0.241** ± 1.043 | **-0.178** ± 0.924 | **-0.146** ± 0.763 |
| S100A1 | **1.628** ± 1.559 | **0.52** ± 1.218 | **-0.723** ± 1.063 | **-0.013** ± 1.464 |
| SMAD3 | **0.108** ± 0.400 | **-0.234** ± 0.711 | **0.3** ± 0.425 | **0.261** ± 0.579 |
| VIM | **-0.836** ± 0.889 | **-0.602** ± 1.134 | **0.805** ± 0.973 | **0.671** ± 0.878 |
| ***Tumor supressor genes*** |  |  |  |  |
| SMARCB1 | **0.023** ± 0.944 | **0.934** ± 0.884 | **0.425** ± 1.005 | **-0.393** ± 0.893 |
| TP53 | **-0.002** ± 0.949 | **0.101** ± 1.026 | **0.703** ± 0.813 | **0.074** ± 0.775 |

First, we analyzed the overall profile expression pattern of each gene among GBM subtypes (Table 2). For the DNA repair and chromatin remodeling genes, such as *ATRX* and *H3F3A*, we observed a tendency to a lower expression level in mesenchymal and an increased expression in proneural compared to neural and classic subtypes. An inverse pattern was observed for *MGMT* with a tendency to be up-regulated in mesenchymal and down-regulated in proneural subtypes with respect to neural and classic subtypes. The *IDH1* gene showed a different pattern for the classical subtype, as it is highly expressed compared to neural, proneural, and mesenchymal subtypes. No changes for *TERT* expression were found.

Among the cytoskeleton and cellular proliferation genes, the most substantial differences among subtypes are for *EGFR*, with a general tendency to be up-regulated in the classical proliferative subtype and down-regulated in the proneural and mesenchymal subtypes. Another tyrosine kinase growth factor, *FLT1*, did not show differences in expression among GBM subtypes. The downstream effectors for growth factors *PIK3CA* and *SMAD3* showed up-regulation and down-regulation, respectively, for the proneural subtype; meanwhile, for the classical and mesenchymal subtype *SMAD3* showed a tendency to up-regulation. Another proliferation biomarker, *MKI67*, showed a marked overexpression in the proneural subtype and a tendency to down-regulation in the neural, classical, and mesenchymal subtypes. *NES* and *VIM* appeared to be expressed more in the classic subtype than in other subtypes. Moreover, no relevant changes were observed for *GFAP*, another intermediate filament expressed in neural stem cells, *OLIG2* is up-regulated in the proneural and down-regulated in the mesenchymal subtypes, and *S100A1* was found to be up-regulated in the proneural subtype. For the tumor suppressor genes, *TP53* is clearly down-regulated in the mesenchymal and, to a lesser degree in the classical subtype, but up-regulated in the less proliferative proneural subtype. In contrast, *SMARCB1* showed a different pattern, being overexpressed only in the classical subtype.

We then analyzed the expression patterns of the driver genes clustered into three subgroups of patients according to their age (Tables 3-5). An important observation is that among tumors showing expression of these genes in patients under 30 years, the neural and mesenchymal subtypes were not observed (Table 3). On the other hand, the driver gene expression in the mesenchymal subtype is only present in patients older than 80 years (Supplementary Figure 1).

**Table 3.** Summary of driver gene expression in GBM molecular subtypes in the 10-29 year subgroup with significant *p*-value.

| YOUNG SUBGROUP (10-29 years) | PRONEURAL | CLASSICAL |
| --- | --- | --- |
| ***DNA Repair and Chromatin remodeling genes*** |  |  |
| ATRX | **0.105** ± 1.316 | **-0.105** ± 0.711 |
| BRAF | **0.257** ± 0.466 | **0.034** ± 0.777 |
| H3F3A | **0.375** ± 0.626 | **-0.202** ± 0.619 |
| MGMT | **0.297** ± 0.494 | **0.176** ± 1.782 |
| TERT | **0.066** ± 0.335 | **0.196** ± 0.464 |
| ***Cytoskeleton and celular proliferation genes*** |  |  |
| EGFR | **-3.133** ± 1.228 | **-4.563** ± 2.382 |
| FLT1 | **-0.946** ± 0.61 | **-0.955** ± 0.539 |
| GFAP | **-0.106** ± 0.914 | **0.262** ± 0.113 |
| IDH1 | **-0.679** ± -0.679 | **-0.910** ± 0.204 |
| MKI67 | **0.820** ± 2.157 | **0.572** ± 1.801 |
| NES | **-0.207** ± 1.018 | **0.822** ± 1.167 |
| OLIG2 | **0.998** ± 1.427 | **-1.341 \*** ± 0.859 |
| PIK3CA | **0.058** ± 0.526 | **0.272** ± 0.844 |
| S100A1 | **0.452** ± 1.032 | **0.101** ± 0.807 |
| SMAD3 | **0.104** ± 0.732 | **0.664** ± 0.438 |
| VIM | **-1.127** ± 1.343 | **1.455 \*** ± 0.353 |
| ***Tumor supressor genes*** |  |  |
| SMARCB1 | **0.830** ± 0.653 | **0.653** ± 0.874 |
| TP53 | **0.286** ± 0.993 | **-0.336** ± 1.242 |

For the young subgroup, the samples were determined to belong only to proneural and classical subtypes, and from the 18 genes analyzed, only *OLIG2* and *VIM* showed a differential pattern in gene expression. *OLIG2* is up-regulated in the proneural tumors, according to its role as a differentiation biomarker. Meanwhile, it revealed a down-regulation tendency in the classical subtype. An inverse pattern is observed for *VIM*, which is down-regulated in proneural and up-regulated in the classical subtype. Interestingly, *EGFR* is down-regulated in both subtypes (Table 3).

Among the subgroup of adult patients, the behavior of the *EGFR* gene stands out as it is up-regulated in the classic subtype and down-regulated in proneural and mesenchymal subtypes (Table 4). The same pattern was observed in the elderly subgroup, but with a bigger gap between subtypes (Table 5). Analyses of genes *ATRX, H3F3A, MGMT, MKi67, NES, OLIG2, S100A1, VIM, SMARCB1*, and *TP53* in the adult and elderly patients (Table 4, Table 5), revealed the same pattern in the expression changes among subtypes as observed in the overall analysis.

**Table 4.** Summary of driver gene expression in GBM molecular subtypes in the 30-59 year subgroup with significant *p*-value.

| ADULT SUBGROUP (30-59 years) | NEURAL | PRONEURAL | CLASSICAL | MESENCHYMAL |
| --- | --- | --- | --- | --- |
| ***DNA Repair and Chromatin remodeling genes*** |  |  |  |  |
| ATRX | -0.230 ± 0.904 | 0.279* ± 0.920 | 0.113* ± 0.608 | -0.180 ± 0.583 |
| BRAF | -0.339 ± 0.381 | -0.125 ± 0.392 | -0.122 ± 0.561 | 0.064* ± 0.505 |
| H3F3A | -0.092 ± 0.575 | 0.375** ± 0.446 | -0.121 ± 0.609 | -0.451* ± 0.602 |
| MGMT | 0.696 ± 1.686 | -0.354 ± 1.236 | 0.200 ± 1.339 | 0.786 ± 1.272 |
| TERT | 0.108 ± 0.413 | 0.099 ± 0.402 | 0.357 ± 0.472 | 0.138 ± 0.451 |
| ***Cytoskeleton and celular proliferation genes*** |  |  |  |  |
| EGFR | 0.516 ± 4.050 | -2.693* ± 3.863 | 3.314* ± 4.265 | -1.970 ± 3.678 |
| FLT1 | -0.808 ± 1.085 | -0.637 ± 0.735 | -0.259 ± 1.091 | 0.194* ± 1.220 |
| GFAP | 0.119 ± 0.663 | 0.055 ± 1.018 | 0.249 ± 0.541 | -0.399 ± 1.054 |
| IDH1 | 0.125 ± 0.586 | -0.054 ± 0.833 | 0.781** ± 1.065 | -0.115 ± 0.889 |
| MKI67 | -1.094 ± 1.453 | 1.065*** ± 1.551 | -0.052* ± 1.278 | -0.061* ± 0.938 |
| NES | -0.913 ± 1.170 | -0.198* ± 0.700 | 1.380** ± 0.884 | -0.151* ± 0.941 |
| OLIG2 | 0.098 ± 0.786 | 1.396*** ± 1.021 | -0.029 ± 1.083 | -1.581*** ± 1.063 |
| PIK3CA | -1.073 ± 0.550 | 0.415*** ± 1.229 | -0.370** ± 0.785 | -0.328** ± 0.625 |
| S100A1 | 1.289 ± 1.270 | 0.467* ± 1.287 | -0.674*** ± 0.930 | 0.127** ± 1.516 |
| SMAD3 | 0.166 ± 0.463 | -0.207 ± 0.838 | 0.271 ± 0.431 | 0.198 ± 0.542 |
| VIM | -0.929 ± 0.927 | -0.369 ± 1.083 | 0.564*** ± 0.850 | 0.778*** ± 0.944 |
| ***Tumor supressor genes*** |  |  |  |  |
| SMARCB1 | 0.171 ± 0.902 | 1.008* ± 0.951 | 0.311 ± 0.998 | -0.297 ± 0.862 |
| TP53 | -0.087 ± 0.992 | 0.194 ± 1.162 | 0.829** ± 0.654 | -0.124 ± 0.809 |

**Table 5.**
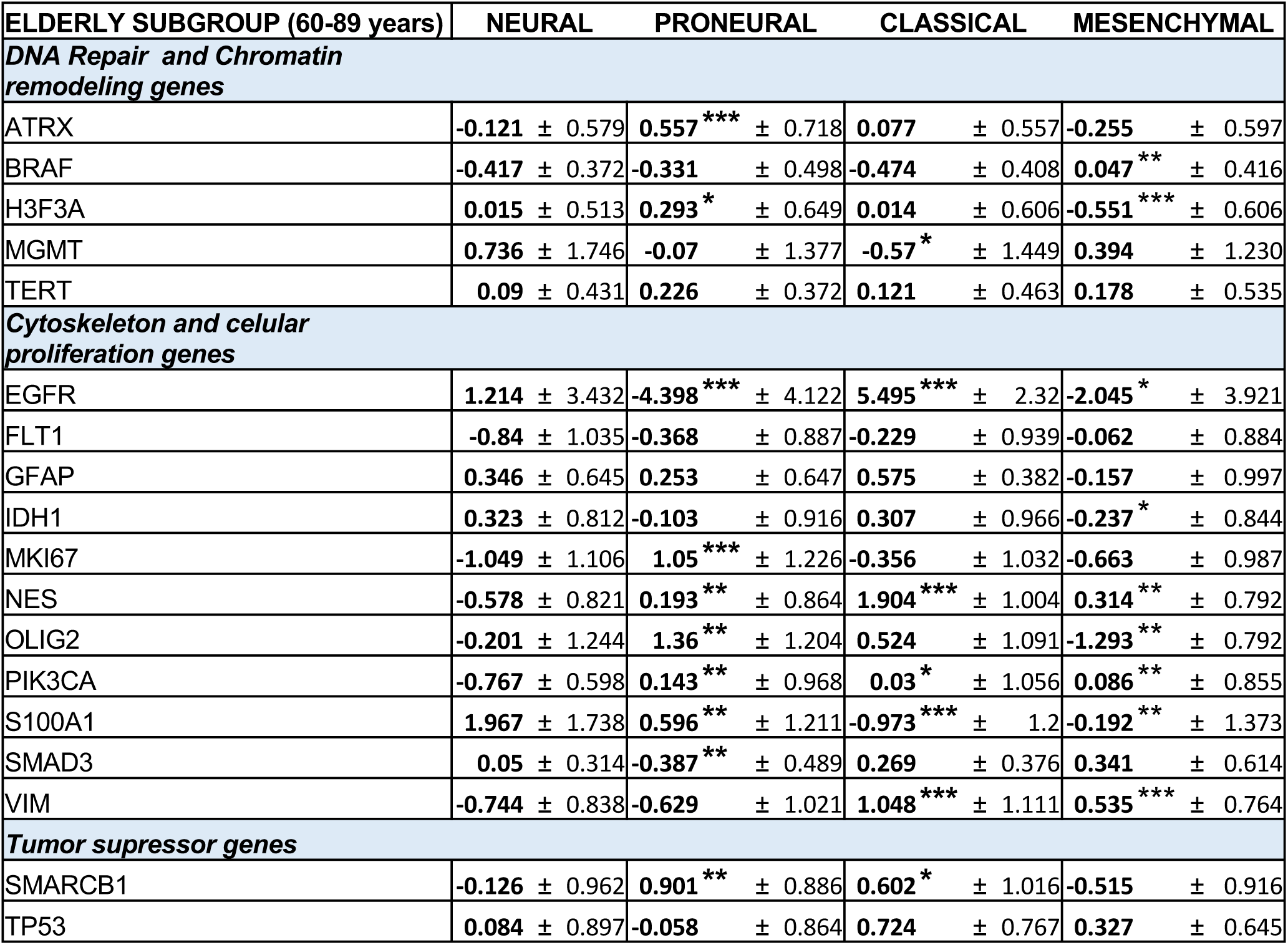
Summary of driver gene expression in GBM molecular subtypes in the 60-89 year subgroup with significant *p*-value.

To analyze the variation of these biomarkers at different stages of life in each subtype, we selected the genes with the most remarkable differential expression pattern. The most common GBM biomarker, *EGFR* gene, showed a remarkable up-regulation in the classic subtype from adult to elderly subgroups, while it was down-regulated in the young subgroup. An inverse pattern is observed in the proneural subtype, with progressive down-regulation from young to adult and elderly subgroups (Figure 1a). For *BRAF*, a differential pattern was observed only in the proneural subtype, being up-regulated in tumors in young patients and down-regulated in elderly patients (Figure 1b). *OLIG2* had a remarkable differential pattern in the classical subtype, in which it is down-regulated in young patients and shows an up-regulation in elderly patients (Figure 1c). *IDH1* expression varies in the classical subtype, being down-regulated in young patients and up-regulated in both adult and elderly patients (Figure 1d). Finally, *GFAP* and *VIM* also showed a variation in expression among the classical subtype samples, but it was of a minimal magnitude (Figure 1e and 1f).

**Figure 1.**
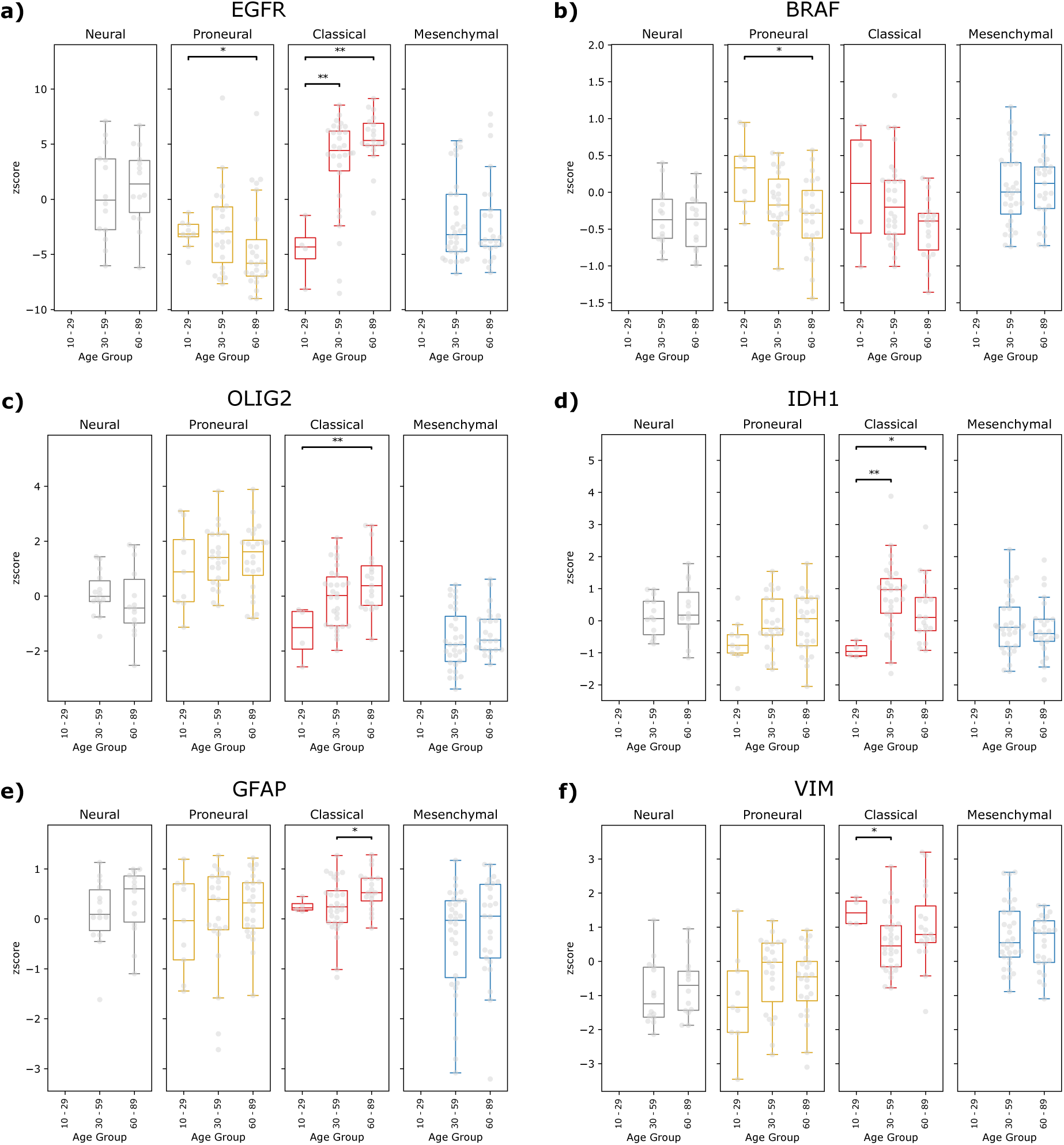
Comparison of driver gene expression profiles among patients grouped by GBM subtype.

Summarizing, the gene expression analysis showed that the altered expression pattern in the mesenchymal subtype include overexpression of *MGMT* that contributes to mutation development, and down-regulation of differentiation biomarkers as *OLIG2* but up-regulation of stemness biomarkers as *VIM*. The altered expression profile in classical subtype includes overexpression of proliferation biomarkers like *EGFR* and stemness biomarkers as *NES* and *VIM*. The expression profile in proneural showed more characteristics of neural progenitor with the up-regulation of *OLIG2* (Table 6).

**Table 6.**
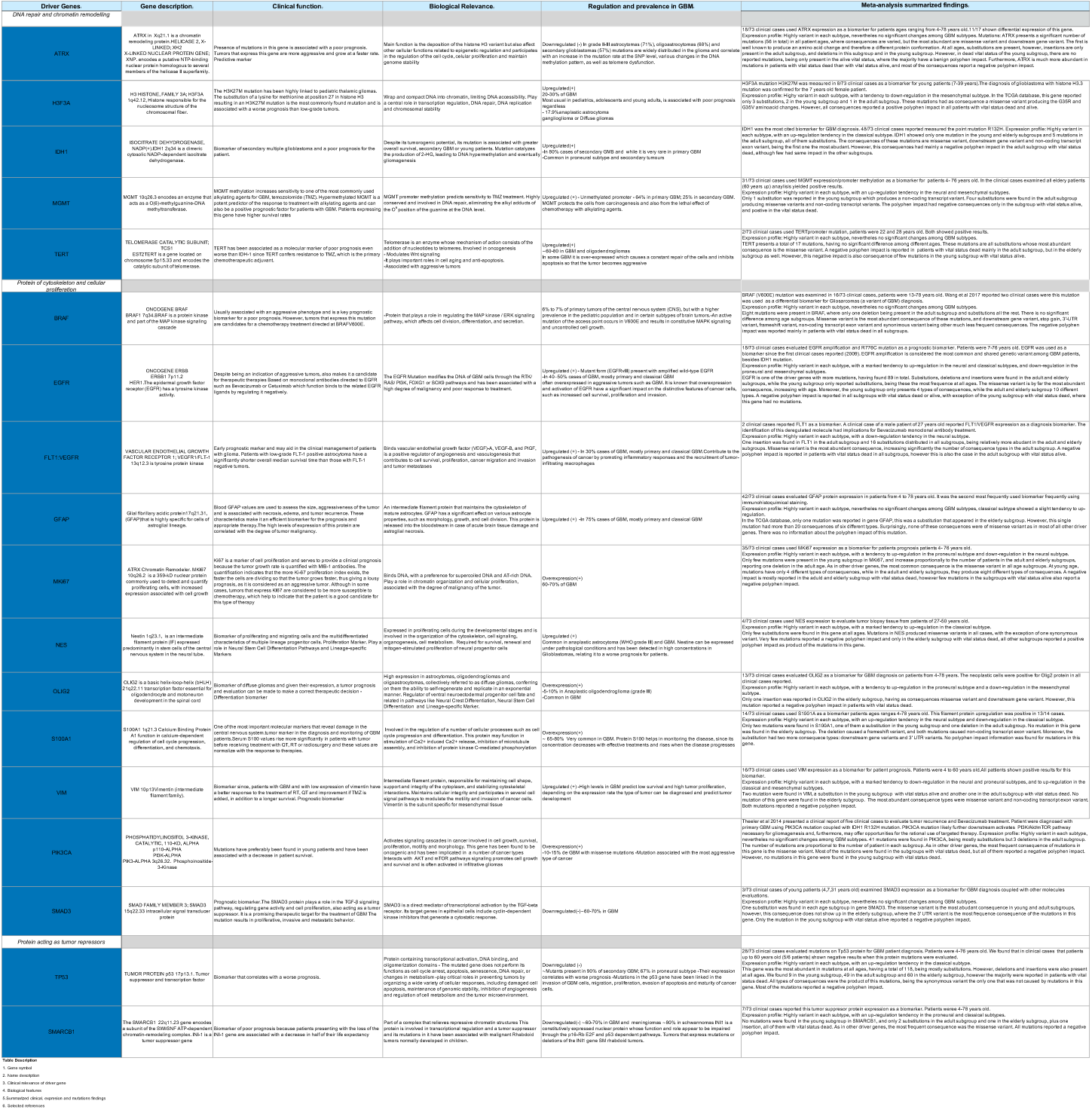
Summary of driver gene features with clinical relevance in GBM diagnosis.

### Somatic Mutation analysis on driver genes

Gene mutation profiling has also served as a biomarker for the diagnosis and treatment of GBM. We used high-throughput data from the TCGA-GBM project and obtained the genomic profiles of a total of 588 clinical cases.

Among the driver genes, clinical cases showed that most frequently affected genes in patients were *TP53* (26%), *EGFR* (22%), *ATRX / PIK3CA* (∼10%), and *IDH1 / MKi67* (∼5%) (Suppl. Figure 2). For an overall view of GBM aberrations, the distribution of the total mutated genes and their DNA changes was determined using the relative percentage of gene mutations according to gene length and number per chromosome. Even though they are not the chromosomes with the highest number of genes, chromosomes 15, 14, and 21 showed the highest percentage of mutations. The lowest percentage was found in chromosomes 18, 10, and Y (Figure 2a). Chromosome 1, which contains the highest number of coding genes (2076), showed a lower percentage of mutations than chromosome 17, which contains less than 60% number of genes (1209). *TP53 (*17p13.1), which suffers from a broad amount of mutations, and *GFAP (*17q21.31), two of the top used genomic markers for their importance due to their genetic alterations, are found in this chromosome (Figure 2b). Among all mutations, 95% substitutions, 3% deletions, and 2% insertions were identified (Figure 2a).

**Figure 2.**
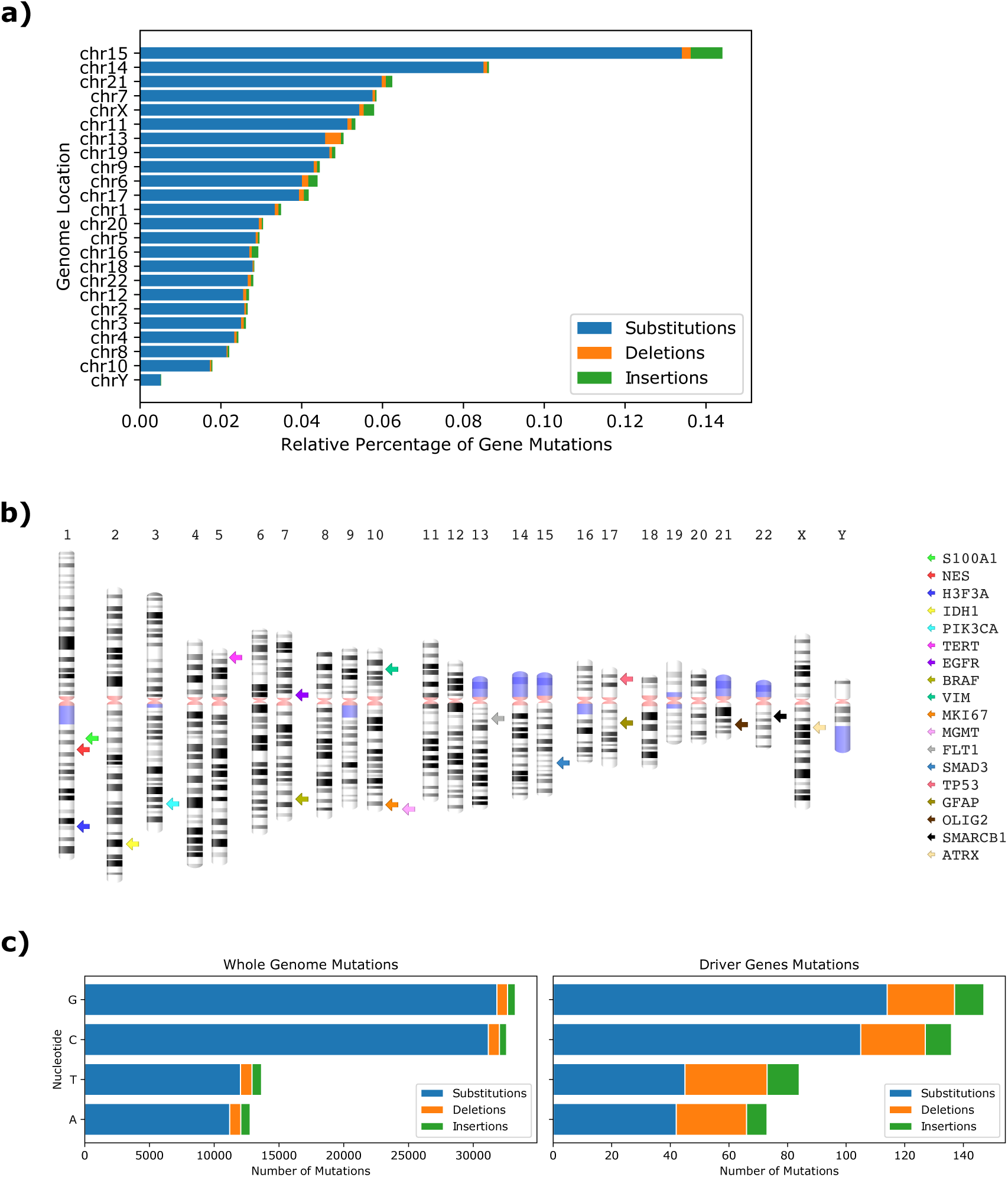
Distribution of the percentage of mutations in genes per chromosome observed in the TCGA-GBM project and the location of their mutations.

A comparison was established to infer whether the total distribution of DNA changes present in all genome-wide mutations was conserved in driver genes, resulting in an affirmative outcome, where DNA changes of nucleotides G-C were the highest (Figure 2c). However, mutations in the driver genes displayed a higher number of deletions and insertions than the whole-genome rates.

The genome location and frequency of gene mutations were determined according to the patient age subgroup. Chromosome 15 showed the highest percentage of mutations in all subgroups. However, some chromosomes, such as 6, 13 and X, showed different patterns according to patient age. Regarding mutation types, substitutions were the highest in all patients, but an increase of deletions and insertions was found according to patient age (Figure 3a). We also observed that mutations in the driver genes reflect the parallel distribution of the genome-wide mutations (Figure 3b), as it is the case in other cancers (16). However, the frequency of mutations varies according to age group, highlighting the different mutational behavior of driver genes in the young subgroup. In particular, *TP53* and *EGFR*, which show to be the most mutated genes in adult and elderly subgroups, are not mutated in the young subgroup, where *ATRX* is the most affected driver gene. Among other DNA repair and chromatin remodeling genes, the mutation frequency behavior of *IDH1*, and *MGMT* increases at 30 years of age and decreases at 60 years (Figure 3b). When analyzing these mutations in more detail, we observed that most of the mutations in all subgroups are substitutions: 91% in young, 80% in adults, and 87% in the elderly.

**Figure 3.**
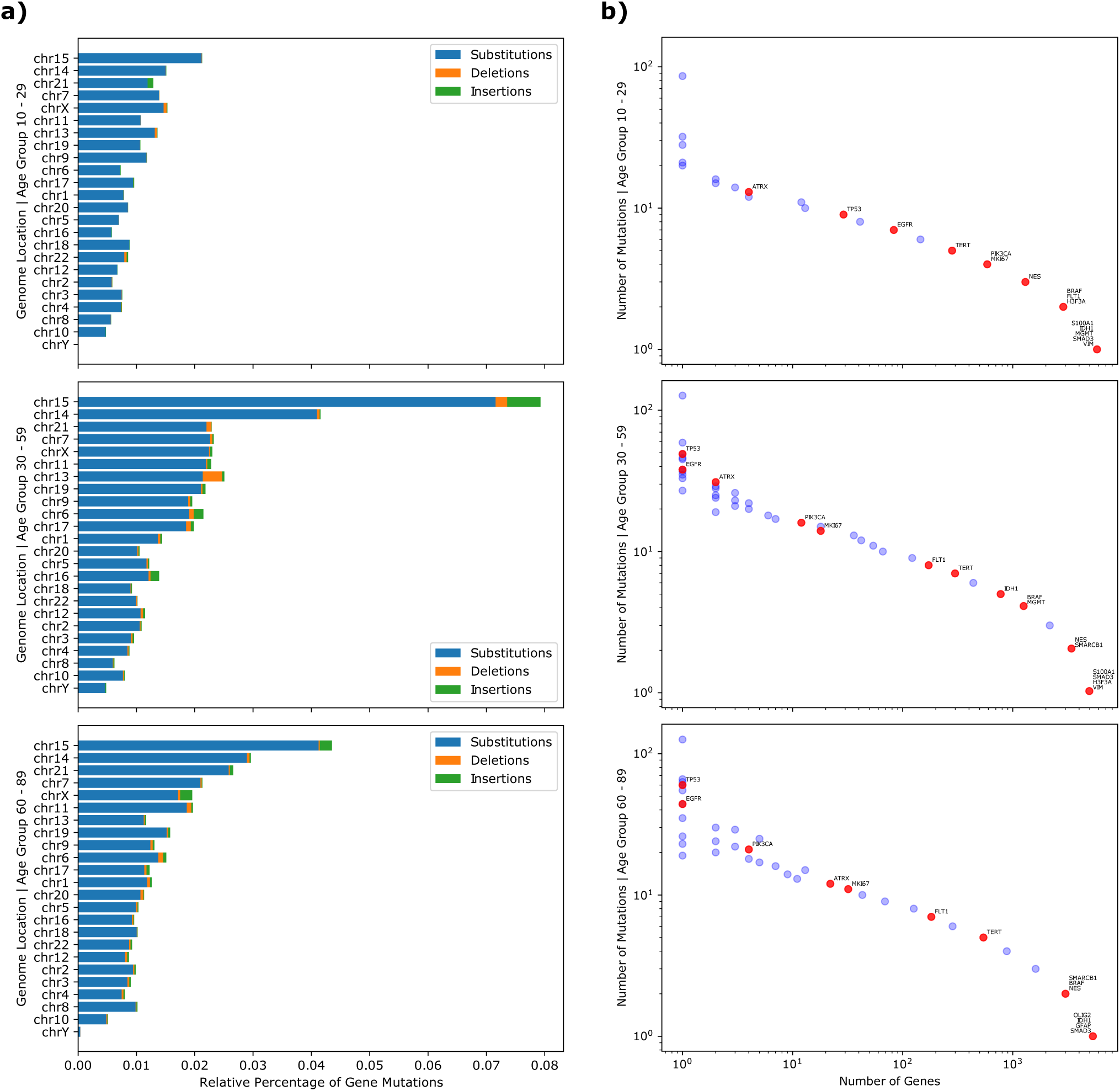
Genome location and percentage of gene mutations according to patient age subgroup.

Summarizing, the *TP53* tumor suppressor gene was found to have the highest frequency of mutations among all patient groups. For *SMARCB1*, another tumor suppressor gene, we found few mutations in adult and elderly subgroups, and none for the young subgroup (Figure 3b).

### Phenotypic consequences of mutations on driver genes

We also studied the phenotypic consequences of each mutation, which can often cause many of them. In the case of *TP53*, for example, a single mutation affects its 27 transcripts, causing consequences of different types. The missense variant consequence appears to be by far the most abundant, representing 47% of all consequences elicited by somatic mutations. Downstream and upstream gene variants, frameshift and intron variants, and stop gain, represent 35% of the consequences caused by mutations, and the remaining percentage is distributed among all other consequences.

Then, we focused on analyzing the biological relevance of mutations on the driver genes. Polymorphism Phenotyping (polyphen) helps to predict the functional significance of an allele replacement from its features by a Naïve Bayes classifier (17). The polyphen impact reported in TCGA is a prediction of a mutation consequence being probably damaging, possibly damaging, or benign. Therefore, we used this data to indicate the possible impact of the consequence types on the function of the proteins encoded by the driver genes. As we found that polyphen impact was mainly reported for the missense variant consequence, we focused on the possible impact of amino acid substitutions.

Driver gene mutations were clustered by patient age and analyzed by their protein phenotype impact values. Among the driver genes, the most affected among all patient samples were *TP53, EGFR, ATRX, PIK3CA, IDH1 and MKi67* (Figure 4). Mutations in the tumor suppressor gene *TP53* represent one of the most common genetic lesions in cancer. In keeping with this, *TP53* was the most affected gene among the driver genes and in the whole genome, increasing abruptly with patient age, as was the case for *EGFR*. In this clinical cohort, among the DNA repair and chromatin remodeling genes, *MGMT*, and *H3F3A* mutations were present only in the young and adult subgroups, with no possible negative impact on their protein functions. In *FLT1, BRAF*, and *MIK167*, the polyphen impact indicates damage in protein functions for the adult subgroup. *NES* and *VIM* mutations are present only among patients below 60 years of age with an unfavorable consequence in protein structure and function. For the *GFAP* and *S1001A* genes, no mutation rates for protein polyphen impact were found. Notably, *OLIG2* mutations with damaging impact consequences were found only in the elderly subgroup.

**Figure 4.**
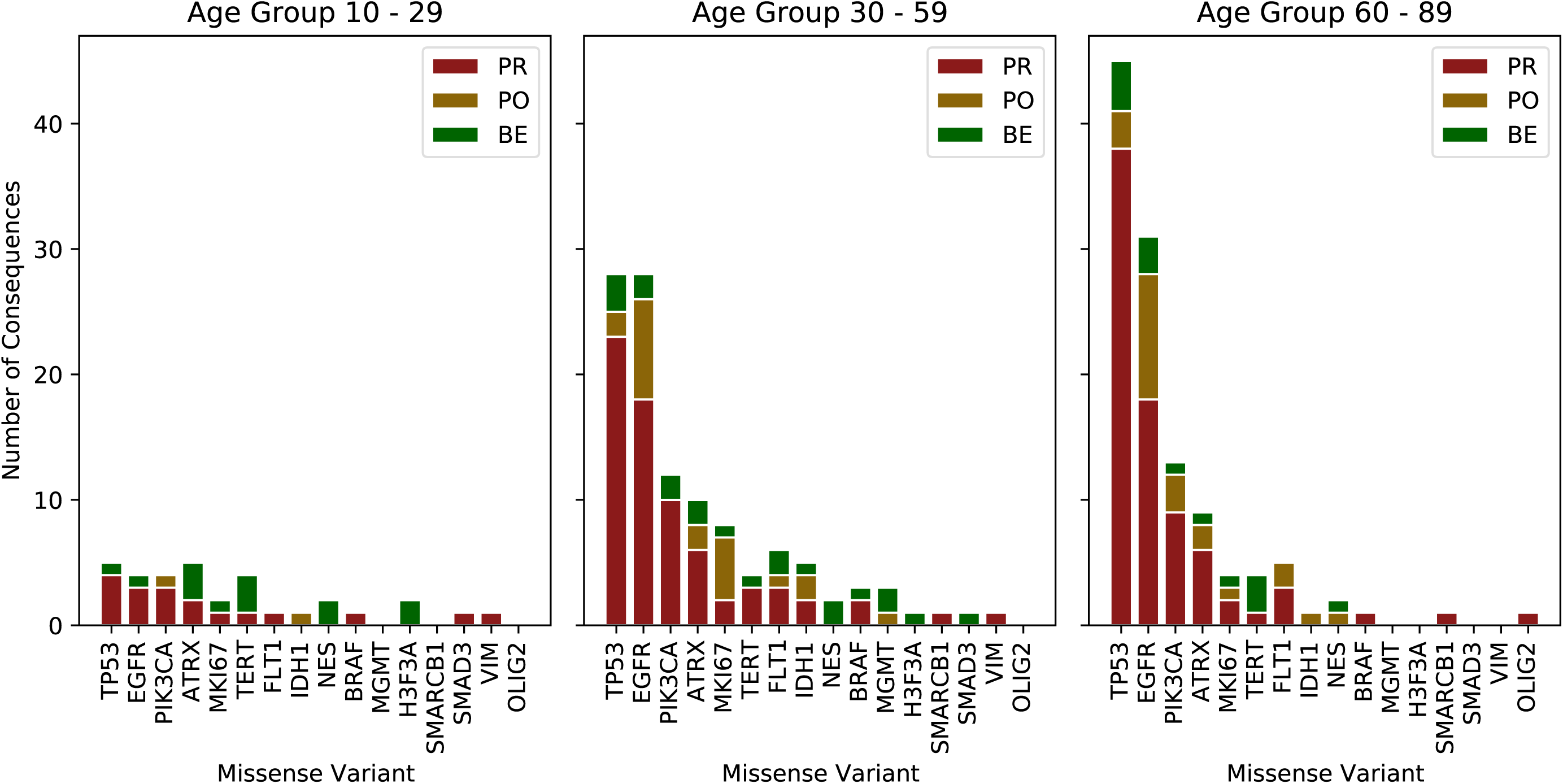
Distribution of protein phenotype impact of mutation-consequences of driver genes grouped by patient age.

### Driver gene biological pathways compared to the highest affected genes in GBM

Functional enrichment analysis was carried out for driver genes and for other genes identified with the worst protein polyphen impact. Driver genes are significantly enriched in hsa:0513 and hsa:0512 for pancreatic and endometrial cancer from the KEGG pathway (-log 10, 9.05> −7.3), and the top GO terms include dsRNA processing, multicellular organism growth, negative regulation of cell differentiation, regulation of DNA metabolic process, and regulation of neuron apoptotic process (-log10 −7.3>-4.80) (Figure 5a). We also observed that the most affected protein phenotypes are functionally enriched in biological processes such as blood circulation, purine containing compound biosynthetic process, cellular response to nitrogen compound, and vascular process in the circulatory system (-log10 −30.02< −22.68) (Figure 5b). The biological pathways enriched were Reactome has R-HSA-382551: Transport of small molecules, (-log10 −46.69), has KEGG has:04022 cGMP PKG signaling pathway, has: in has:0513 and 00071 Fatty acid degradation, and has:00010 Glycolysis/gluconeogenesis pathways (-log10 −39.80 > −16.74) (Figure 5b). Those lesioned genes were linked to seizures, epilepsy, weight loss, pediatric failure to thrive, mental depression, irritation, and vomiting symptoms (-log10 −18< −8.3) (Figure 5b).

**Figure 5.**
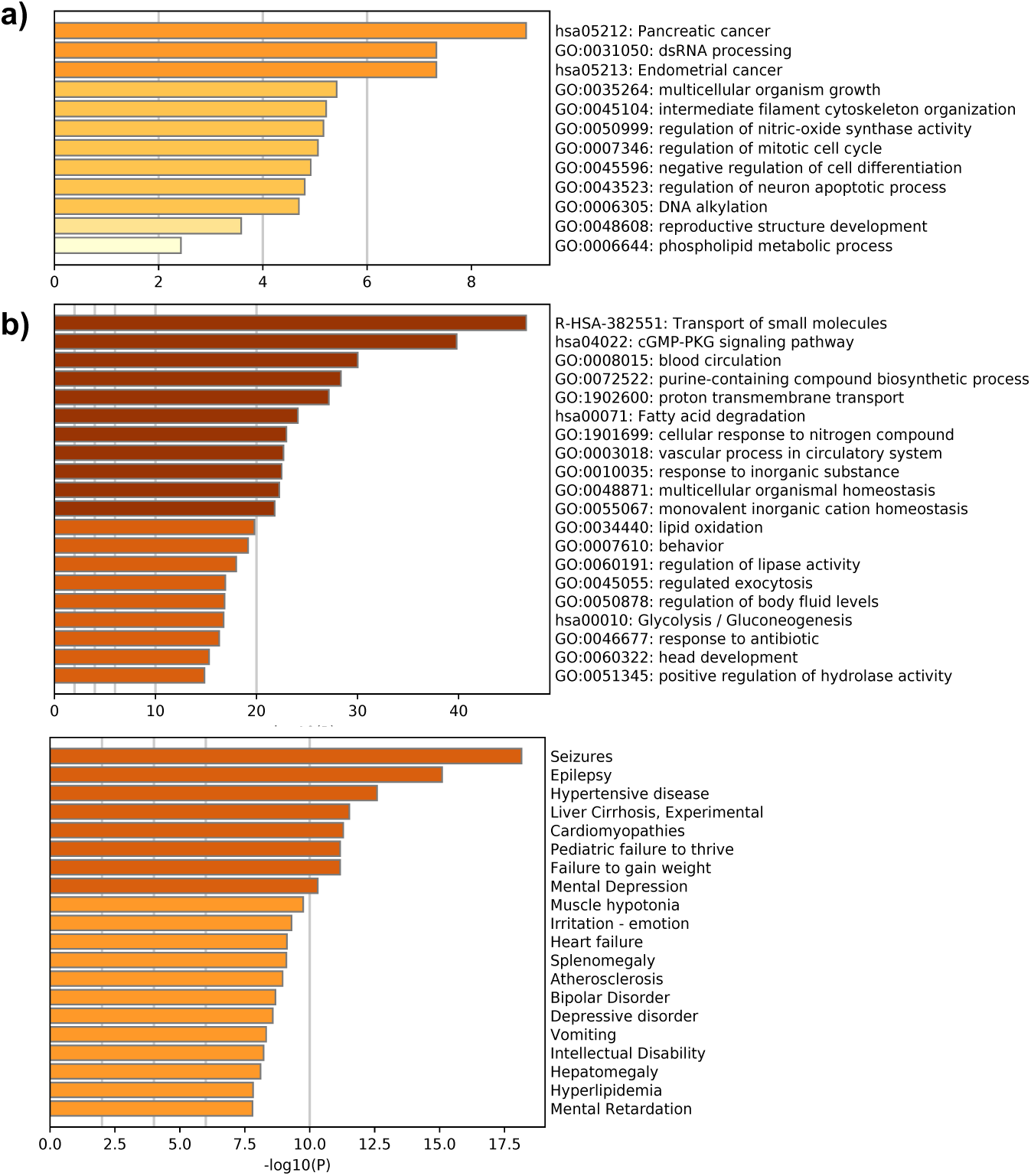
Comparison of driver gene ontology enrichment analysis with the most lesioned genes in the TCGA-GBM project

## Discussion

Current clinical standard methods in neuro-oncology for GBM diagnosis consist of tumor surgery resection and biopsy followed by pathology analysis. We searched literature over the last vicennium and found sixty clinical reports of seventy-three clinical cases in which patient tumor biopsy or fluid sample underwent the analysis of a combination of genomic biomarkers which mainly consisted in *IDH1, GFAP, MKi67*, and *MGMT* coupled in sets with more than two and up to eleven additional markers per sample for diagnosis. Genomic markers were reported for their relevance as measurable indicators of the presence and severity of GBM. Among those genes, the measures on the expression of *ATRX, MGMT, FLT1, GFAP, MKi67, NES, OLIG2, S1001A, VIM, PIK3CA*, as well as the genetic analysis of driver mutation events in *BRAF, H3F3A, TERT, EGFR, IDH1, SMAD3, TP53*, and *SMARCB1* were highlighted from our literature search strategy. We searched among clinical results for a pattern of biomarker behavior in the analyzed samples with unsuccessful results. Aware of inter-tumor molecular heterogeneity as a significant challenge, and due to the remarkable importance of driver genes for the routine clinical role, we delved into their biological behavior. A compendium of summarized findings of driver genes is shown in Table 6.

GBM inter-tumor heterogeneity allows molecular subclassification based on genomic profiling. This is also affected by intra-tumor heterogeneity, originating from two proposed mechanisms, clonal evolution, and cancer stem cells. Clonal evolution is the process by which a single cell undergoes reiterative genetic changes which allows it to evolve and disseminate, forming a tumor (18). In contrast, cancer stem cells in GBM could possess different stemness according to their cellular ontology, being a direct transformation from a normal stem cell or a reprogramming process from a cancer stem cell with less proliferative or differentiation capacity (15). The GBM tumor consists of a core region of high cell proliferation and inflammation, delimited by a margin between the tumor tissue and the normal brain cells, and then the PBZ mainly composed of normal tissue with some infiltrative and isolated tumor cells (14).

Based on a meta-analysis, we herein describe the heterogeneity of GBM at the transcriptional and the genomic levels, with an emphasis on tumorigenesis driver genes currently used in the clinic as genomic markers. Altogether, our results suggest that a combination of these biomarkers would provide a multidimensional approach for a better diagnosis and GBM subtype molecular classification for patient prognosis. Besides, our studies for gene expression and somatic mutations will provide information on the heterogeneity of primary GBM types due to their clinical relevance.

Our meta-analysis from mRNA expression data agrees with previous reports with respect to the mesenchymal subtype. This subtype is characterized by its poor prognosis, stem cell biomarkers, angiogenesis, a prominent radio, and chemoresistance. From the eighteen genes analyzed, we found up-regulation of *MGMT*, which may be related to its own promoter’s unmethylated status frequently observed in this GBM subtype and related to Temozolomide treatment resistance and short patient survival (19). In our analysis, this expression profile was conserved during adult and elderly life stages.

Furthermore, the down-regulation of *ATRX, H3F3A*, and *EGFR* was observed. *ATRX* encodes an adaptor protein that contributes to the Methyl-CpG binding protein 2 (*MeCP2*) mediated pericentric heterochromatin organization, which is very important for neural differentiation (20); thus, down-regulation of this gene might be expected in a less differentiated subtype with more stemness such as the mesenchymal GBM subtype cells. The opposite, up-regulated behavior, was observed in the proneural subtype, which has less stemness and more characteristics of a differentiated cell. Another chromatin remodeler, *H3F3A*, whose driver mutations HK27M and G34R induce dysfunction of Polycomb repressive complex 2 (*PRC2*) and dramatic alterations of gene expression (21, 22), may contribute to high alterations in profile expression for mesenchymal GBM subtype. *EGFR*, which is perhaps one of the best-characterized molecules in primary GBM (23), showed a down-regulation in mesenchymal and proneural subtypes, but a clear up-regulation in the classical GBM subtype. This behavior is conserved across all age groups and strikingly marked for the elderly population. This expression profile could be dependent on mesenchymal GBM increased mutation rates, which may play a feedback role downregulating *EGFR* gene expression. Co-existence of mutations in critical molecules from downstream *EGFR* signaling such as Ras or *PTEN*, which maintain active signaling without a ligand to the receptor, could play a role as an alternative mechanism.

We observed other genes with striking profile expression, including *NES, VIM*, and *TP53*, with up-regulation behavior. *NES* and *VIM* encode the intermediate filament proteins Nestin and Vimentin. Vimentin is expressed mainly in mesenchymal cell types, while Nestin mainly in neural stem and progenitor cells in the central nervous system (24). These proteins function not only as part of the cytoskeleton, but also impact several key cellular processes such as proliferation, death, migration, and invasiveness (24). Our analysis showed that *VIM* is up-regulated in both mesenchymal and classical GBM subtypes and *NES* only in the classical subtype. This pattern may be related to the ontogenesis of these tumors and suggest the transition state for classical GBM to a possible mesenchymal GBM, but with a neural stem cell marker remaining.

The proneural GBM subtype showed up-regulation of *MKI67* and *OLIG2. MKI67* encodes the DNA binding protein Ki-67 and is widely used as a proliferation marker as it participates in chromosome motility and chromatin organization during the cell cycle (25). *OLIG2* encodes a central nervous system transcription factor that plays an essential role in the proliferation of oligodendrocyte precursors and their differentiation (26). *OLIG2* also showed down-regulation in classical and mesenchymal GBM subtypes. Therefore, these expression patterns support the idea that the proneural GBM subtype arises from central nervous system progenitors with fewer stemness properties but with proliferative capacity.

Our analysis in the expression profile for the eighteen driver genes supports the GBM ontogenesis hypothesis from Celiku *et al*. 2014 (15), which proposes that proneural subtypes can be generated from neural progenitors, and these cells may gain somatic mutations to become classical and consecutively mesenchymal subtypes. It is also possible that classical or mesenchymal subtypes originate from central nervous system progenitors with high stemness.

In this study, we found that all driver genes have reported mutations in GBM patients. However, genes that are significantly mutated and that display multiple biological consequences include *TP53, ATRX, PIK3CA*, and *EGFR*. Abnormalities of the *TP53* gene have been the most extensively investigated genetic variations found in more than 50% of human tumors (28). Contrary to other reports where *TP53* mutations are more related to pediatric tumors (29), we found an increasing behavior from the young to elderly subgroups. The same behavior is observed for genes *ATRX, PIK3CA*, and *EGFR*. However, *TP53* and *EGFR* were found to be the most mutated genes in adult and elderly subgroups, while these mutational behavior changes in the young subgroup, where *ATRX* is the most affected gene (Figure 4b).

Impairment of the DNA repair process is expected to increase the overall frequency of mutations and, hence, the likelihood of cancer-causing mutations. In comparison to other studies in which *ATRX* is mutated only rarely in adult primary GBM, but frequently found in younger adults with lower-grade glioma (WHO grade II/III)(30), we found a high frequency at 30 years that decreases in elderly patients. Similar behavior was identified for *IDH1* and *MGMT* DNA repair and chromatin remodeling genes.

Additionally, *NES* and *VIM* mutations were absent in the elderly subgroup and are present only in patients below 60 years-old with an unfavorable consequence in protein structure and function. In contrast, *OLIG2* mutations with negative impact consequences were found only in the elderly patient subgroup.

Some driver mutations on key genes have been pivotal for the diagnosis and prognosis of GBM patients. We focused particularly on the effects of mutations with non-synonymous changes, also called missense mutations, which alter the codons so that they specify different amino acids during protein synthesis (Suppl. Figure. 3), and carried out a comparison of GO enriched terms of the selected driver genes with the genes identified with a higher probability of damaging consequences. We found that highly affected pathways such as blood circulation and vascular processes in the circulatory system are consistent with alterations in angiogenesis in GBM. We also identified a link of the lesioned proteins to seizures, epilepsy, weight loss, pediatric failure to thrive, mental depression, irritation, and vomiting, among other symptoms which are in agreement with those reported in the clinical cases reviewed.

Efforts have been made for the identification of relevant biomarkers to assess GBM progression by targeting genes with the highest density of missense mutations. For example, tumors with the *BRAF* V600E mutation tend to be more severe. This somatic mutation prevents *Braf* protein from controlling cell proliferation (Suppl. Figure 3), which has been reported in the TCGA database, appearing at all ages but more frequently in elderly patients.

*TP53* mutations were predominantly point mutations, which lead to amino acid substitutions in the DNA binding domain (DBD). The substitution of arginine residues within the DBD, such as R175, R248, and R273, was reported in other studies and was also found in the GBM patients (31). However, this was not the most abundant amino acid substitution, being G105R, S127Y, P152S, and V157G, examples of some amino acid changes abundantly reported in the TCGA cohort.

The most cited biomarker for diagnosis *IDH1* R132H has also been reported in the TCGA database as a mutation in all age subgroups with a negative polyphen impact (3). On the other hand, the H3K27M mutation that has been highly linked to pediatric thalamic gliomas and is associated with a worse prognosis than low-grade tumors was not found in the TCGA cohort, which is the case of other biomarkers used in clinical studies, such as H3G34R, H3G34N, *EGFR* R776C, and the *TERT* promoter mutations C228T and C250T (21, 22, 30).

To better understand the behavior of mutations in young patients, we briefly analyzed genes that are involved in GBM with the worst polyphen impact consequences and analyzed the transcription factors that regulate them. Our results showed that the young subgroup behaves differently, as genes that are mutated are regulated by different TFs. Moreover, the TFs that regulate genes with mutations in the young subgroup share almost no TFs with adult and elderly subgroups. This might explain why the young subgroup has a divergent behavior in comparison with the other subgroups. On the other hand, the adult and elderly subgroups share most of the biological pathways, while microtubule cytoskeleton organization, regulation of microtubule based process, adenylate cyclase-inhibiting G protein-coupled glutamate receptor signaling among others are GO terms unique for the adult subgroup, whilst protein-protein interactions at synapses, regulation of cyclase activity and carbohydrate digestion and absorption are unique functional terms for the eldery subgroup. In particular, genes with mutations with a negative polyphen impact in the 10-29-year-old subgroup share fewer identities with the 60-89 year-old subgroup (Suppl. Figure 4).

It is surprising that among all the TCGA data reported for GBM, several mutations that are defined as biomarkers could not be found. The absence of a clearly defined and concordant pattern between clinical, transcriptomics, and mutational dynamics studies, support the idea of outstanding heterogeneity in GBM. Despite the high abundance of somatic aberrations in GBM tumors, only a select few have been associated with clinical relevance and are currently used as biomarkers. No single mutation has been identified to trigger a particular type of GBM tumor. The intra and inter tumor heterogeneity of GBM has revealed its “multiforme” nature not only at its morphologic and phenotypic levels but also on its genotype.

Furthermore, the relationship between genetic alterations and gene expression at the mRNA level is not always linear. The interplay between distant genetic interactions and epigenetic changes also have a significant impact on the expression of specific genes. Hence, the selection of the most commonly mutated and amplified genes as therapeutic targets may not be sufficient. Our results showed that the link of genomic markers and profile expression with their phenotypic alterations is more complex than previously thought. With this analysis, however, we expect to contribute to the construction of a panel of driver genes to better delineate the intra- and inter-tumor heterogeneity for a more accurate diagnosis. To achieve this objective, it is crucial to analyze the raw data for other key molecules involved in the mechanisms that drive the balance between proliferation and differentiation in the stem and cell precursors for the central nervous system.

Currently expression levels of *ATRX, MGMT, FLT1, GFAP, MKi67, NES, OLIG2, S1001A, VIM*, and *PIK3CA* are used in the clinic for patient GBM diagnosis and prognosis. Our results suggest that the biomarker set integrated by *ATRX, H3F3A, TP53, EGFR, NES, VIM, Mki67, MGMT* and *OLIG2* genes could be a strong combination to determine the GBM molecular subtype (Figure 6). For example, the mesenchymal subtype, known as the most aggressive GBM, showed overexpression of *MGMT* and the repression of *ATRX, H3F3A, TP53*, and *EGFR*. On the other hand, while overexpression of *EGFR, NES, VIM*, and *TP53* was characteristic of the proliferative or classical subtype, if there is overexpression of *Mki67* and *OLIG2*, the prognosis could be more favorable owing to their association with the less aggressive proneural subtype (Figure 6). Further clinical trials with patient samples for expression analysis using the abovementioned biomarkers could provide confirmatory evidence for their clinical potential.

## Conclusions

GBM is a highly heterogeneous cancer that consists of multiple molecular alterations. Despite the vertiginous advances in the clinical medical area, the prognosis of patients continues to be unfavorable, with an average of overall survival of less than one year. The differential molecular characteristics of histologically similar tumors make it difficult to predict clinical outcomes and select optimal treatment strategies. Given the heterogeneity of GBM and the multitude of factors that influence disease progression, general clinical characteristics are insufficient to predict individual prognosis and survival accurately. In clinical routine, a combination of biomarkers is necessary for patient differential diagnosis and prognosis being *IDH1, GFAP, Mki67*, and *MGMT* the most reported ones. The inter-tumor molecular heterogeneity remains the hardest challenge in neuro-oncology practice. In our study, the expression profiles of those markers revealed a consistent link with the glioma progression model for tumor ontogenesis, supporting that GBM tumors display a unique behavior and that “personalized” treatment must be required for each molecular subtype. Our results suggest that a set of the following biomarkers *ATRX, EGFR, H3F3A, MGMT, Mki67, NES, TP53, OLIG2 and VIM* genes could be a strong combination to determine the GBM molecular subtype for patient prognosis. Notably, the frequency of mutations varies according to age group, highlighting the different mutational behavior of driver genes in the young subgroup. In particular, *TP53* and *EGFR*, which show to be the most mutated genes in the adult and elderly subgroups, are not mutated in the young subgroup, in which *ATRX* is the most affected driver gene. Besides, a unique distribution of somatic mutations was found for the young and adult populations, particularly for genes related to DNA repair and chromatin remodeling *ATRX, MGMT, and IDH1*. We also highlighted differential patient age regulatory and biological pathway behaviors that could serve as a basis for further analysis in the journey of the development of improved therapy for patients suffering from this disease.

## Data Availability

Python scripts (https://github.com/kap8416/GBM-META-ANALYSIS-OF-DRIVER-GENES)
mRNA expression analysis using data from the Glioblastoma BioDiscovery Portal (GBM-BioDP) https://gbm-biodp.nci.nih.gov
Genomic data for the eighteen genes previously identified as molecular markers was downloaded from the NIH website https://portal.gdc.cancer.gov/

## Acknowledgments

K.A.P thanks to the CABANA program for bioinformatics training. For technical support, we thank Luis Alberto Aguilar Bautista, Alejandro de León Cuevas, Carlos Sair Flores Bautista, and Jair García of the Laboratorio Nacional de Visualización Científica Avanzada (LAVIS).

---

**Supplementary Figure 1.**
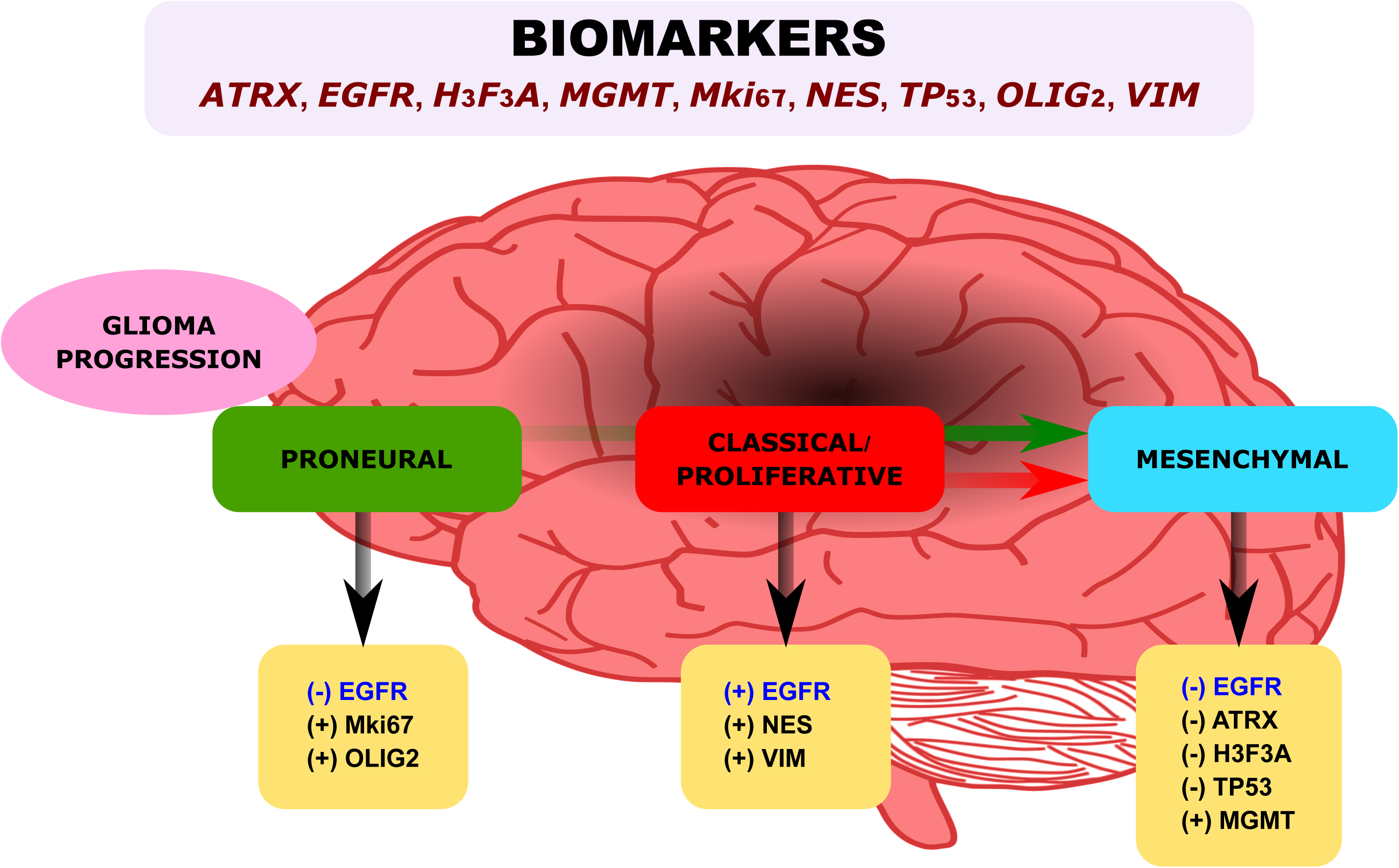
Distribution of GBM molecular subtypes across age groups.

**Supplementary Figure 2.**
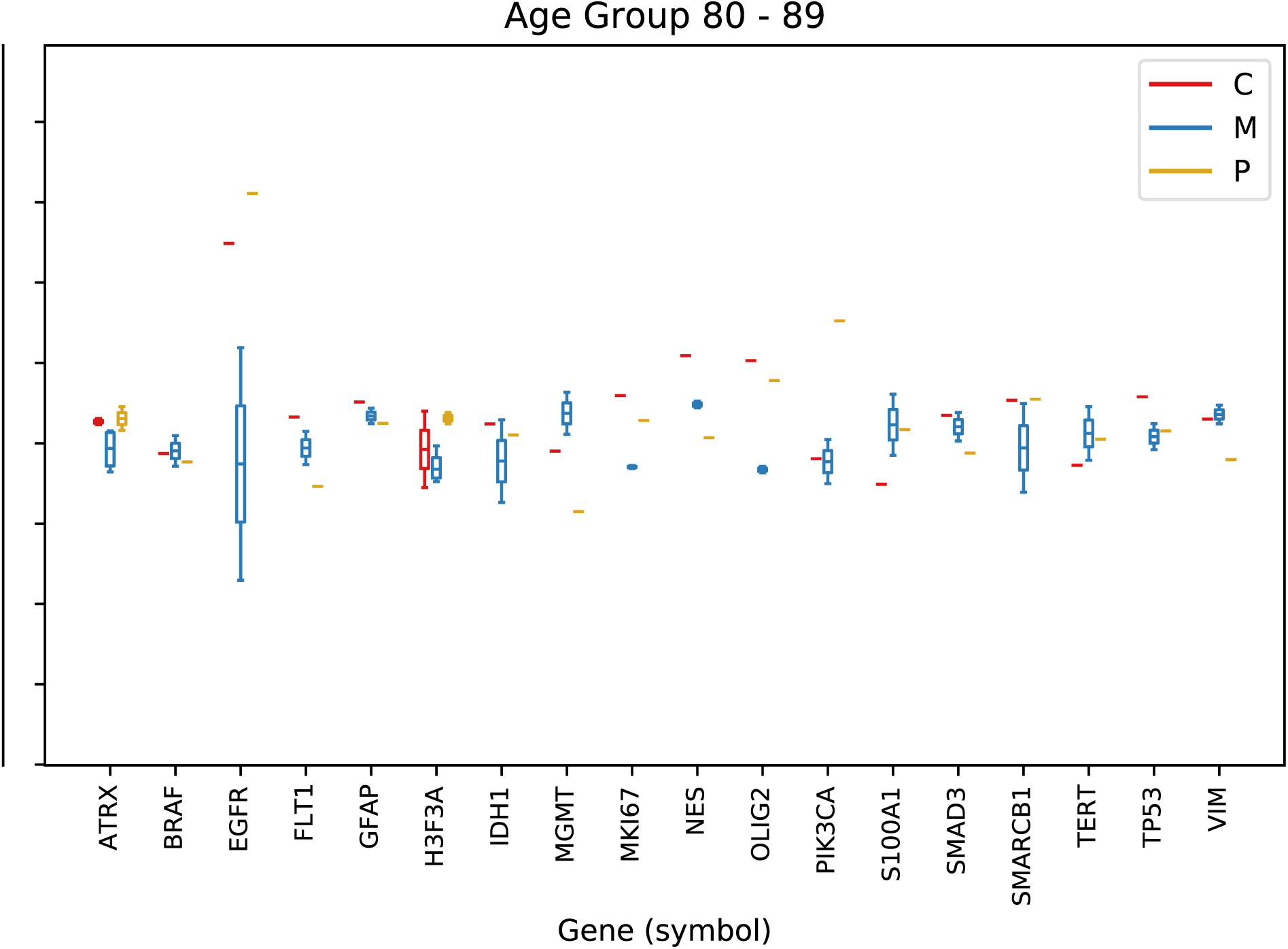
Frequence of mutations in the driver genes.

**Supplementary Figure 3.**
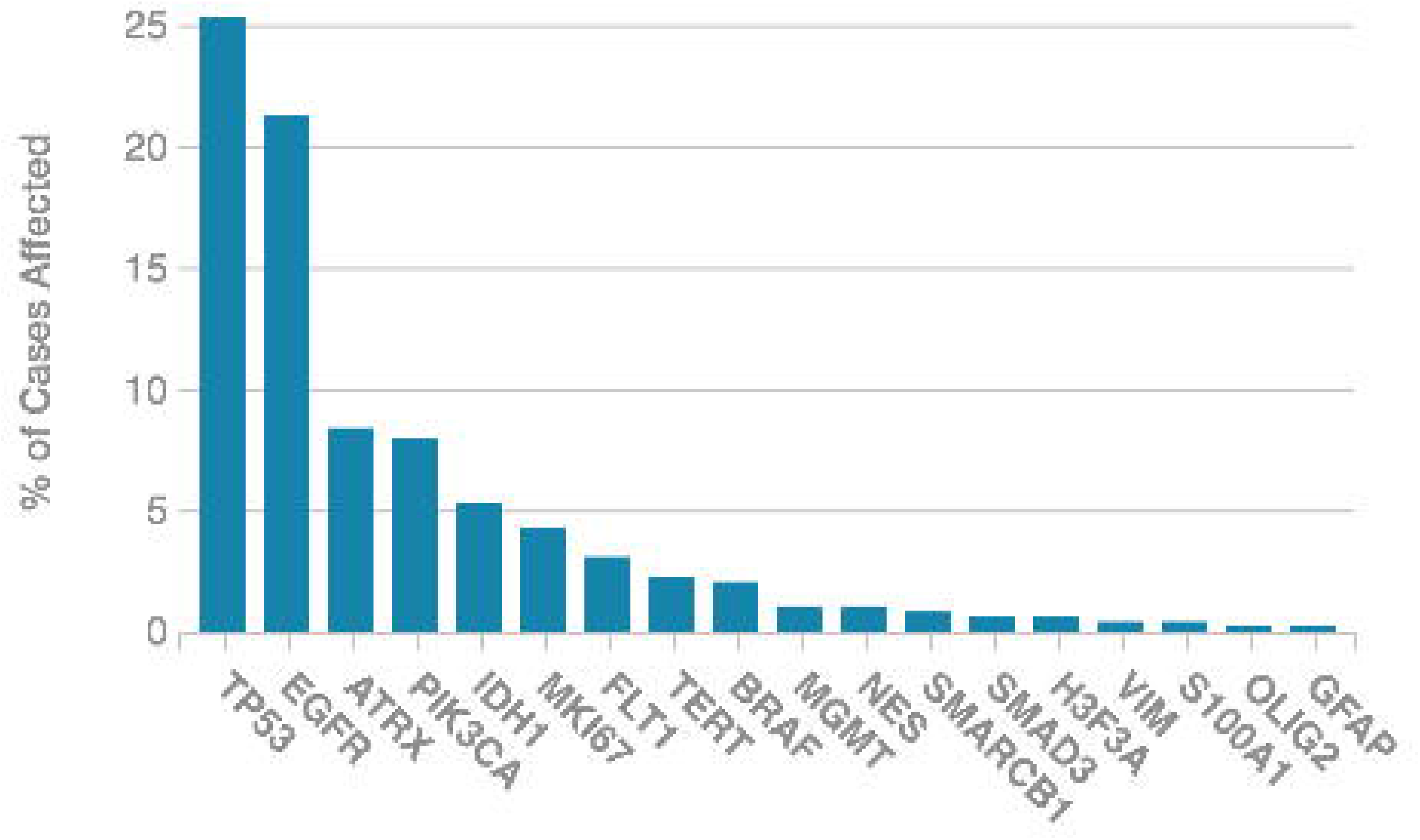
Distribution of amino acid changes in proteins of selected driver genes.

**Figure 4.**
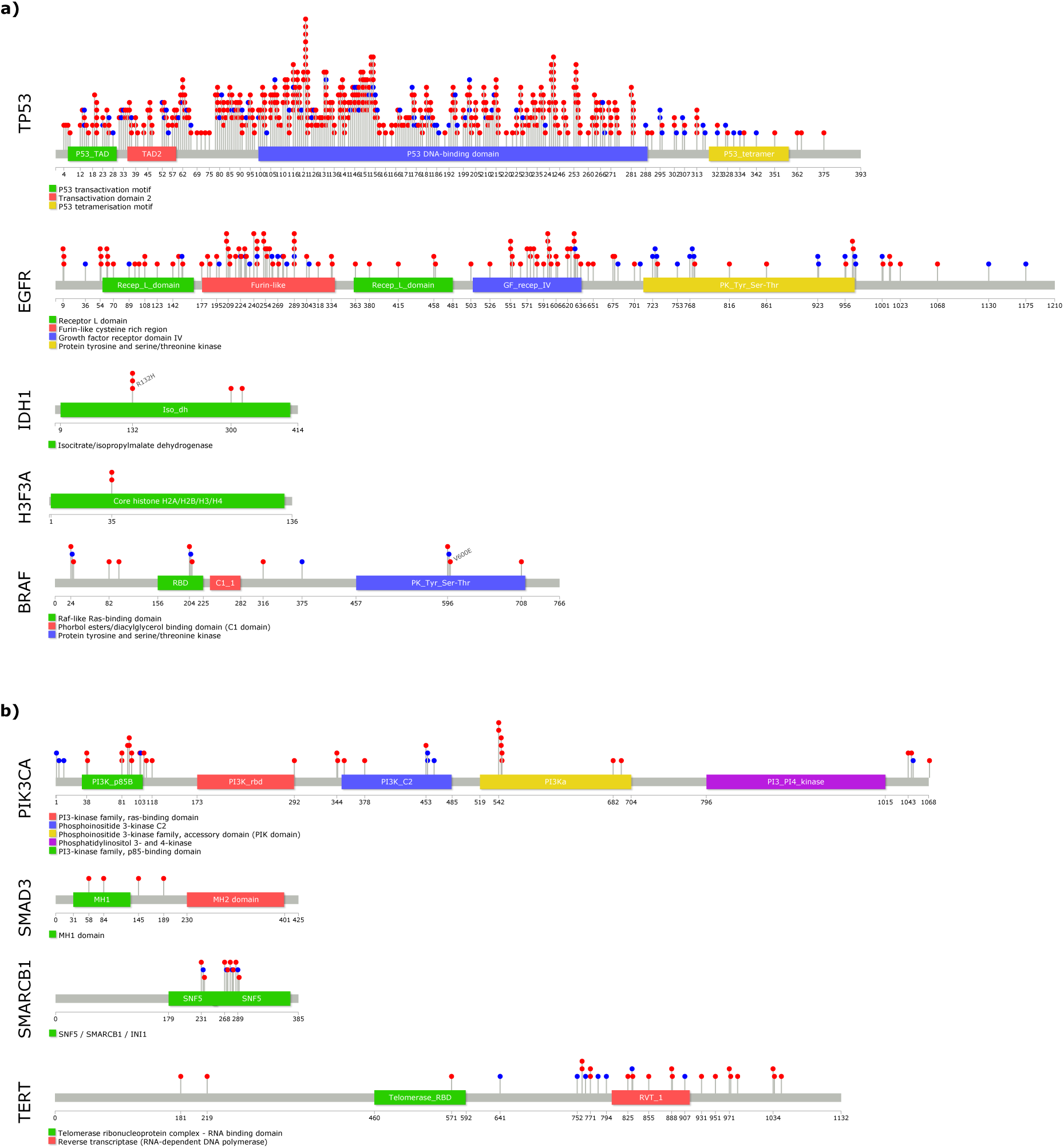
Enrichment analysis of the most affected genes in polyphen impact grouped by patient age.

**Supplemental File 1.**
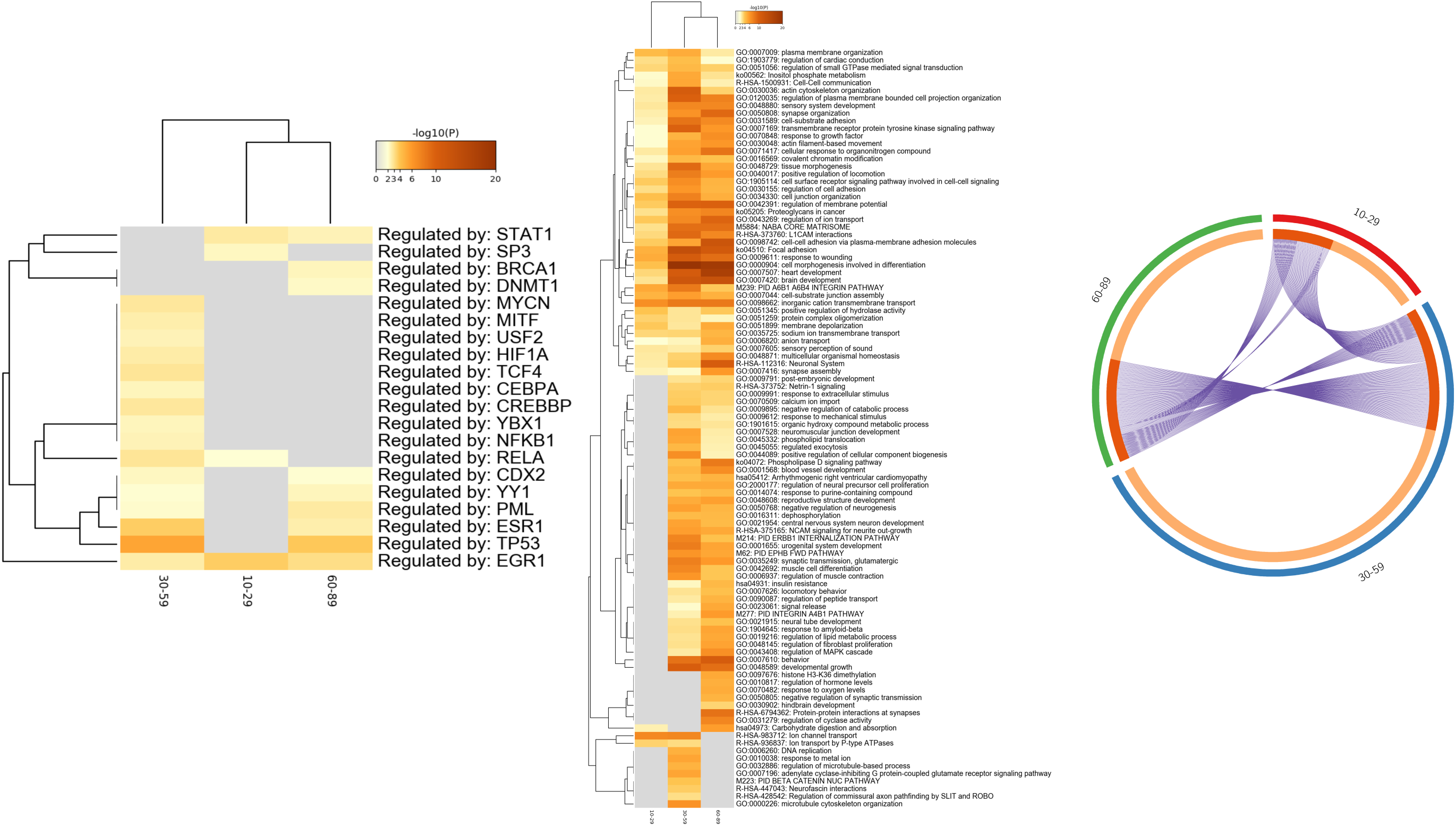
PRISMA Statement of clinical report meta-analysis.

## References

1. Tykocki T, Eltayeb M. Ten-year survival in glioblastoma. A systematic review. J Clin Neurosci. 2018;54:7–13.

2. Patel AP, Tirosh I, Trombetta JJ, Shalek AK, Gillespie SM, Wakimoto H, et al. Single-cell RNA-seq highlights intratumoral heterogeneity in primary glioblastoma. Science. 2014;344(6190):1396–401.

3. Ohgaki H, Kleihues P. The definition of Primary and Secondary Glioblastoma.Clin Cancer Res. 2013; 19(4) 764–772.

4. Mansouri A, Karamchandani J, Das S. Molecular Genetics of Secondary Glioblastoma. In: De Vleeschouwer S, editor. Glioblastoma. Brisbane (AU)2017.

5. Li R, Li H, Yan W, Yang P, Bao Z, Zhang C, et al. Genetic and clinical characteristics of primary and secondary glioblastoma are associated with differential molecular subtype distribution. Oncotarget. 2015;6(9):7318–24.

6. Han J, Puri RK. Analysis of the cancer genome atlas (TCGA) database identifies an inverse relationship between interleukin-13 receptor alpha1 and alpha2 gene expression and poor prognosis and drug resistance in subjects with glioblastoma multiforme. J Neurooncol. 2018;136(3):463–74.

7. Alifieris C, Trafalis DT. Glioblastoma multiforme: Pathogenesis and treatment. Pharmacol Ther. 2015;152:63–82.

8. Szopa W, Burley TA, Kramer-Marek G, Kaspera W. Diagnostic and Therapeutic Biomarkers in Glioblastoma: Current Status and Future Perspectives. Biomed Res Int. 2017;2017:8013575.

9. Huse JT, Holland EC. Targeting brain cancer: advances in the molecular pathology of malignant glioma and medulloblastoma. Nat Rev Cancer. 2010;10(5):319–31.

10. Vizcaino MA, Shah S, Eberhart CG, Rodriguez FJ. Clinicopathologic implications of NF1 gene alterations in diffuse gliomas. Hum Pathol. 2015;46(9):1323–30.

11. Padfield E, Ellis HP, Kurian KM. Current Therapeutic Advances Targeting EGFR and EGFRvIII in Glioblastoma. Front Oncol. 2015;5:5.

12. Verhaak RG, Hoadley KA, Purdom E, Wang V, Qi Y, Wilkerson MD, et al. Integrated genomic analysis identifies clinically relevant subtypes of glioblastoma characterized by abnormalities in PDGFRA, IDH1, EGFR, and NF1. Cancer Cell. 2010;17(1):98–110.

13. Sonoda Y. Clinical impact of revisions to the WHO classification of diffuse gliomas and associated future problems. Int J Clin Oncol. 2020;25(6):1004–9.

14. Aubry M, de Tayrac M, Etcheverry A, Clavreul A, Saikali S, Menei P, et al. From the core to beyond the margin: a genomic picture of glioblastoma intratumor heterogeneity. Oncotarget. 2015;6(14):12094–109.

15. Celiku O, Johnson S, Zhao S, Camphausen K, Shankavaram U. Visualizing molecular profiles of glioblastoma with GBM-BioDP. PLoS One. 2014;9(7):e101239.

16. Romero-Arias JR, Ramírez-Santiago G, Velasco-Hernández JX, Ohm L, Hernández-Rosales M. Model for breast cancer diversity and spatial heterogeneity. American Physical Society. 2018;98(3):032401.

17. Adzhubei I, Jordan DM, Sunyaev SR. Predicting functional effect of human missense mutations using PolyPhen-2. Curr Protoc Hum Genet. 2013;Chapter 7:Unit7 20.

18. Greaves M, Maley CC. Clonal evolution in cancer. Nature. 2012;481(7381):306–13.

19. Kirstein A, Schmid TE, Combs SE. The Role of miRNA for the Treatment of MGMT Unmethylated Glioblastoma Multiforme. Cancers (Basel). 2020;12(5).

20. Marano D, Fioriniello S, Fiorillo F, Gibbons RJ, D’Esposito M, Della Ragione F. ATRX Contributes to MeCP2-Mediated Pericentric Heterochromatin Organization during Neural Differentiation. Int J Mol Sci. 2019;20(21).

21. Cantero D, Mollejo M, Sepulveda JM, D’Haene N, Gutierrez-Guaman MJ, Rodriguez de Lope A, et al. TP53, ATRX alterations, and low tumor mutation load feature IDH-wildtype giant cell glioblastoma despite exceptional ultra-mutated tumors. Neurooncol Adv. 2020;2(1):vdz059.

22. Duan H, Hu JL, Chen ZH, Li JH, He ZQ, Wang ZN, et al. Assessment of circulating tumor DNA in cerebrospinal fluid by whole exome sequencing to detect genomic alterations of glioblastoma. Chin Med J (Engl). 2020;133(12):1415–21.

23. Alexandru O, Horescu C, Sevastre AS, Cioc CE, Baloi C, Oprita A, et al. Receptor tyrosine kinase targeting in glioblastoma: performance, limitations and future approaches. Contemp Oncol (Pozn). 2020;24(1):55–66.

24. Sharma P, Alsharif S, Fallatah A, Chung BM. Intermediate Filaments as Effectors of Cancer Development and Metastasis: A Focus on Keratins, Vimentin, and Nestin. Cells. 2019;8(5).

25. Menon SS, Guruvayoorappan C, Sakthivel KM, Rasmi RR. Ki-67 protein as a tumour proliferation marker. Clin Chim Acta. 2019;491:39–45.

26. Kosty J, Lu F, Kupp R, Mehta S, Lu QR. Harnessing OLIG2 function in tumorigenicity and plasticity to target malignant gliomas. Cell Cycle. 2017;16(18):1654–60.

27. Lee JH, Lee JE, Kahng JY, Kim SH, Park JS, Yoon SJ, et al. Human glioblastoma arises from subventricular zone cells with low-level driver mutations. Nature. 2018;560(7717):243–7.

28. Leroy B, Anderson M, Soussi T. TP53 mutations in human cancer: database reassessment and prospects for the next decade. Hum Mutat. 2014;35(6):672–88.

29. Pollack IF, Finkelstein SD, Woods J, Burnham J, Holmes EJ, Hamilton RL, et al. Expression of p53 and prognosis in children with malignant gliomas. N Engl J Med. 2002;346(6):420–7.

30. Suzuki H, Aoki K, Chiba K, Sato Y, Shiozawa Y, Shiraishi Y, et al. Mutational landscape and clonal architecture in grade II and III gliomas. Nat Genet. 2015;47(5):458–68.

31. Ham SW, Jeon HY, Jin X, Kim EJ, Kim JK, Shin YJ, et al. TP53 gain-of-function mutation promotes inflammation in glioblastoma. Cell Death Differ. 2019;26(3):409–25.

