## Supplemental File 1 for "GLIOBLASTOMA MULTIFORME: A META-ANALYSIS OF DRIVER GENES, CURRENT DIAGNOSIS, AND TUMOR HETEROGENEITY"

### Supporting Information for Meta-analysis

**Authors:** Gabriel Emilio Herrera-Oropeza[1,2], Carla Angulo-Rojo[3], Santos Alberto Gástelum-López[4], Alfredo Varela-Echavarría[1], Maribel Hernández-Rosales[5]*, and Katia Aviña-Padilla[1]*.

### PRISMA 2009 Checklist

| Section/Topic | # | Checklist Item | Reported on Page # |
|---|---|---|---|
| **TITLE** | | | |
| Title | 1 | Identify the report as a systematic review, meta-analysis, or both. | Page1 |
| **ABSTRACT** | | | |
| Structured summary | 2 | Provide a structured summary including, as applicable: background; objectives; data sources; study eligibility criteria, participants, and interventions; study appraisal and synthesis methods; results; limitations; conclusions and implications of key findings; systematic review registration number. | Page 2-3 |
| **INTRODUCTION** | | | |
| Rationale | 3 | Describe the rationale for the review in the context of what is already known. | Page 4 |
| Objectives | 4 | Provide an explicit statement of questions being addressed with reference to participants, interventions, comparisons, outcomes, and study design (PICOS). | Page 4 |
| **METHODS** | | | |
| Protocol and registration | 5 | Indicate if a review protocol exists, if and where it can be accessed (e.g., Web address), and, if available, provide registration information including registration number. | |
| Eligibility criteria | 6 | Specify study characteristics (e.g., PICOS, length of follow-up) and report characteristics (e.g., years considered, language, publication status) used as criteria for eligibility, giving rationale. | Page 5-6 |
| Information sources | 7 | Describe all information sources (e.g., databases with dates of coverage, contact with study authors to identify additional studies) in the search and date last searched. | Page 5 |
| Search | 8 | Present full electronic search strategy for at least one database, including any limits used, such that it could be repeated. | Page 5 |
| Study selection | 9 | State the process for selecting studies (i.e., screening, eligibility, included in systematic review, and, if applicable, included in the meta-analysis). | Page 5-6 |
| Data collection process | 10 | Describe method of data extraction from reports (e.g., piloted forms, independently, in duplicate) and any processes for obtaining and confirming data from investigators. | Page 6-7 |
| Data items | 11 | List and define all variables for which data were sought (e.g., PICOS, funding sources) and any assumptions and simplifications made. | Table 1 |
| Risk of bias in individual studies | 12 | Describe methods used for assessing risk of bias of individual studies (including specification of whether this was done at the study or outcome level), and how this information is to be used in any data synthesis. | Page 7-8 |
| Summary measures | 13 | State the principal summary measures (e.g., risk ratio, difference in means). | Page 7-8 |
| Synthesis of results | 14 | Describe the methods of handling data and combining results of studies, if done, including measures of consistency (e.g., $I^2$) for each meta-analysis. | Page 8 |

#### PRISMA 2009 Checklist

| Section/Topic | # | Checklist Item | Reported on Page # |
|---|---|---|---|
| Risk of bias across studies | 15 | Specify any assessment of risk of bias that may affect the cumulative evidence (e.g., publication bias, selective reporting within studies). | Page 11 |
| Additional analyses | 16 | Describe methods of additional analyses (e.g., sensitivity or subgroup analyses, meta-regression), if done, indicating which were pre-specified. | Page 11 |
| **RESULTS** | | | |
| Study selection | 17 | Give numbers of studies screened, assessed for eligibility, and included in the review, with reasons for exclusions at each stage, ideally with a flow diagram. | Page 8-9 Figure 1 |
| Study characteristics | 18 | For each study, present characteristics for which data were extracted (e.g., study size, PICOS, follow-up period) and provide the citations. | Page 8-9 Table 1 |
| Risk of bias within studies | 19 | Present data on risk of bias of each study and, if available, any outcome level assessment (see item 12). | Page 11 Figure 6 |
| Results of individual studies | 20 | For all outcomes considered (benefits or harms), present, for each study: (a) simple summary data for each intervention group (b) effect estimates and confidence intervals, ideally with a forest plot. | Table 1 |
| Synthesis of results | 21 | Present the main results of the review. If meta-analyses done, include for each, confidence intervals and measures of consistency. | Page 5 |
| Risk of bias across studies | 22 | Present results of any assessment of risk of bias across studies (see Item 15). | Page 11 Figure 6 |
| Additional analysis | 23 | Give results of additional analyses, if done (e.g., sensitivity or subgroup analyses, meta-regression [see Item 16]). | Page 11 Table 3 Figure 7 |
| **DISCUSSION** | | | |
| Summary of evidence | 24 | Summarize the main findings including the strength of evidence for each main outcome; consider their relevance to key groups (e.g., healthcare providers, users, and policy makers). | Page 12-13 |
| Limitations | 25 | Discuss limitations at study and outcome level (e.g., risk of bias), and at review-level (e.g., incomplete retrieval of identified research, reporting bias). | Page 16-17 |
| Conclusions | 26 | Provide a general interpretation of the results in the context of other evidence, and implications for future research. | Page17 |
| **FUNDING** | | | |

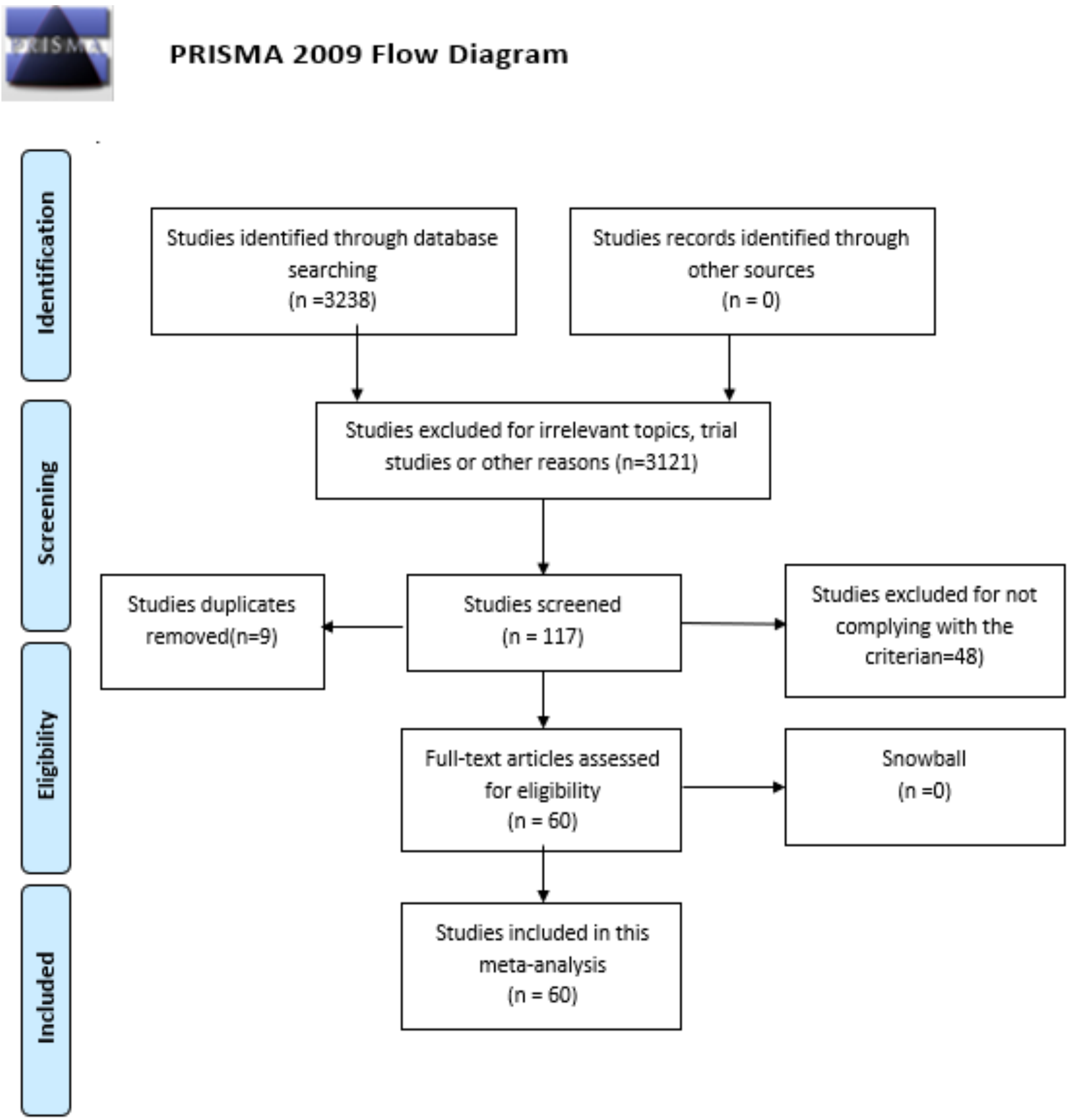


**Figure 1. PRISMA flow diagram.**

### PRISMA flow diagram

### .Methodology

- The aim of the clinical cases meta-analysis was to extract the clinical features, and the top used genomic markers for GBM diagnosis. The literature search was performed in July 2020 for all the cases reported from June 2000 to June 2020.

### Search strategy, and selection criteria

- **1.-Identification.**
- First, we searched in BVS, Chrocane, Karger and PubMed databases, with predefined search terms. (June 2000-June 2020). Data mining key words were: 1 (“*Glioblastoma”*), 2 (”*Clinical”*) and 3 (“*Case or Cases or Report*”). In total, search engines returned 3,238 results.

- Example., in the PubMed.gov

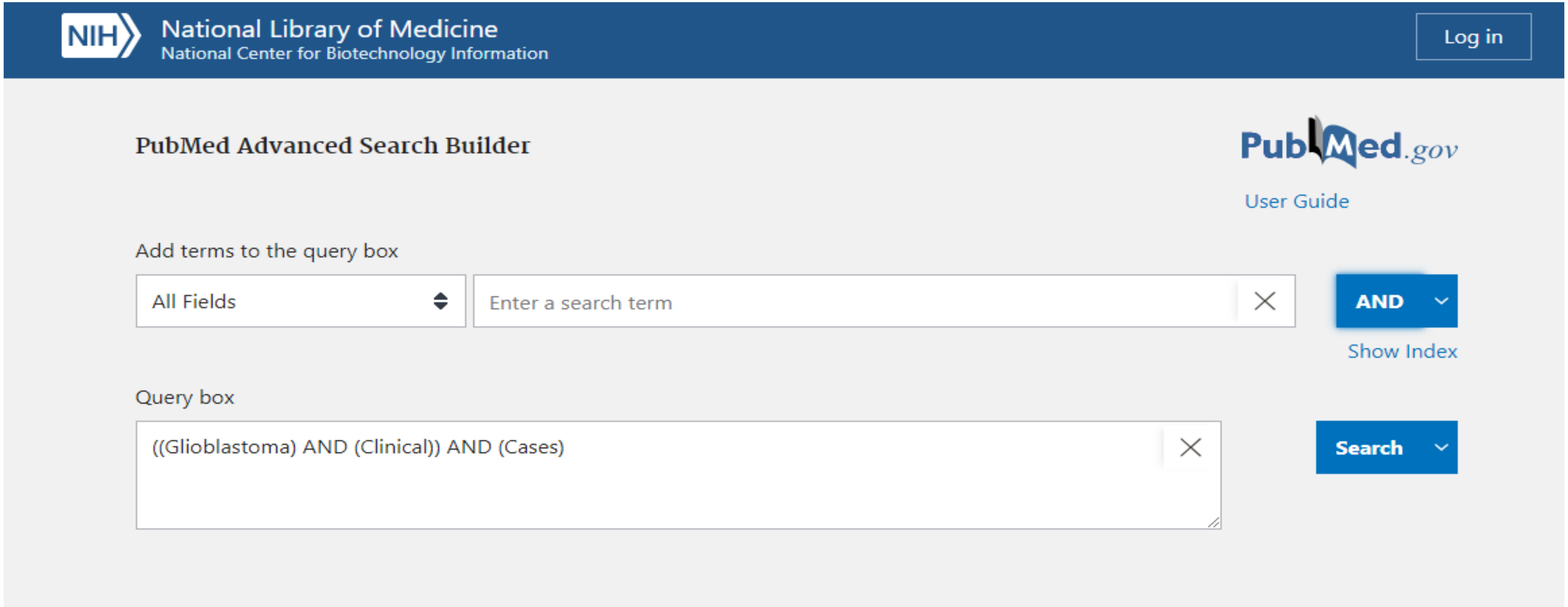

### Search strategy, and selection criteria

- **1.-Identification**
- The search was conducted in July 2020 and focused on studies published in the last vicennium (June 2000 – June 2020).

**Glioblastoma** Multiforme, Diagnosis and Tr

Batash R, Asna N, Schaffer P, Francis N, Schaffer M.

Curr Med Chem. 2017;24(27):3002-3009. doi: 10.2174/0[illegible]

PMID: 28521700 Review.

BACKGROUND: **Glioblastoma** multiforme (GBM) is the [illegible]

with an incidence of 3.19 **cases** per 100,000 person year[illegible]

year survival rate of 4-5%, and only a 26-33% survival ra[illegible]

Genetic Alterations in Gliosarcoma and Giar[illegible]

Oh JE, Ohta T, Nonoguchi N, Satomi K, Capper D, Piersci[illegible]

Antonelli M, Kleihues P, Giangaspero F, Ohgaki H.

Brain Pathol. 2016 Jul;26(4):517-22. doi: 10.1111/bpa.12[illegible]

PMID: 26443480

### 1. A sample of search strategy

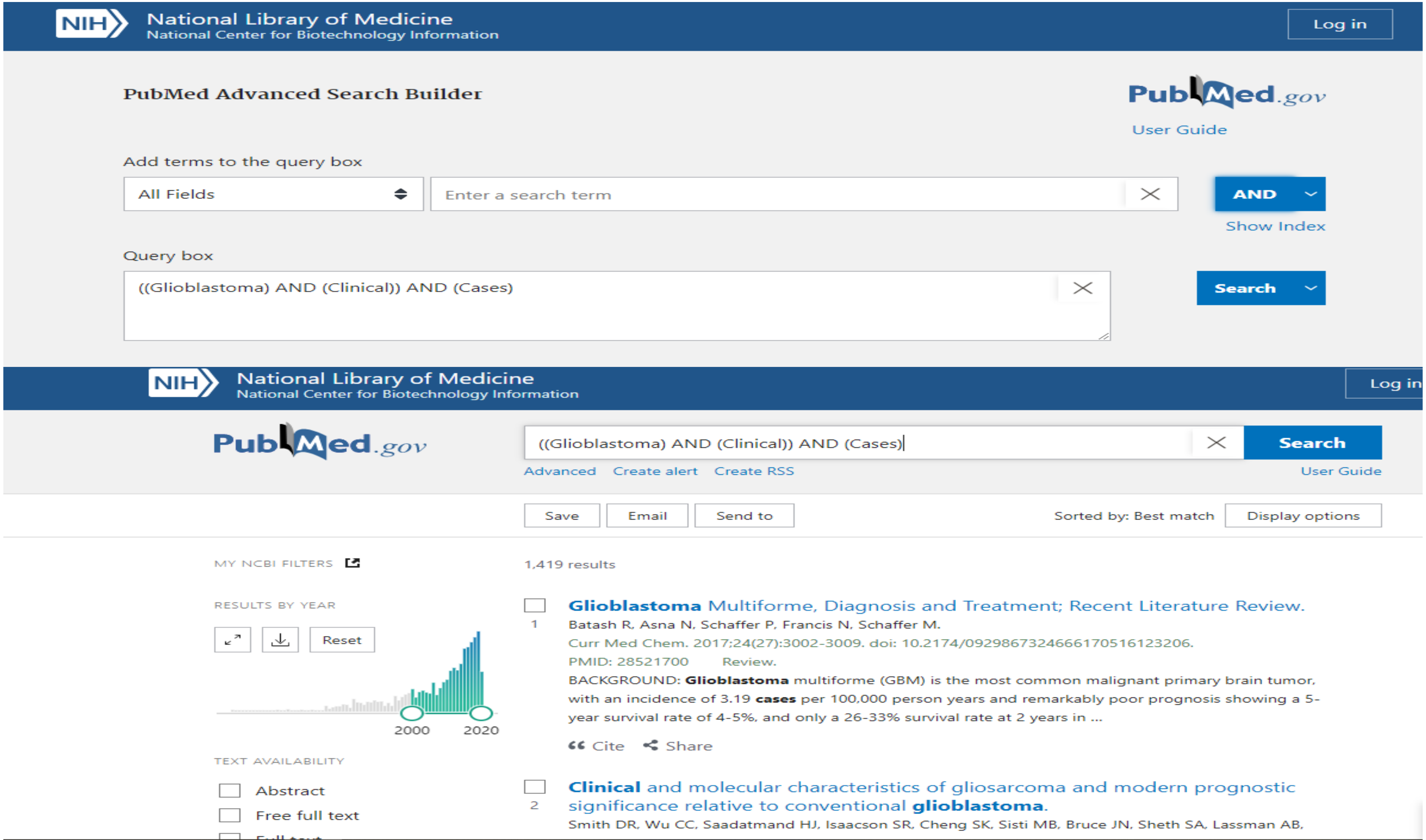

### 2.-Screening

- Titles and abstracts were reviewed, studies that did not meet the requirements were excluded, such as (1) *Glioblastoma*, (2) *Clinical,* and (3) *Cases* or *Report*. The minimum requirements for the consideration of the cases were a brief description of the patient, a confirmatory study of GBM and a follow-up of at least 1 year.
- Studies that did not meet the criteria were excluded. Taking into account these basic inclusion criteria, 3121 publications were excluded after selection of the title and summary and 9 duplicated studies were removed.

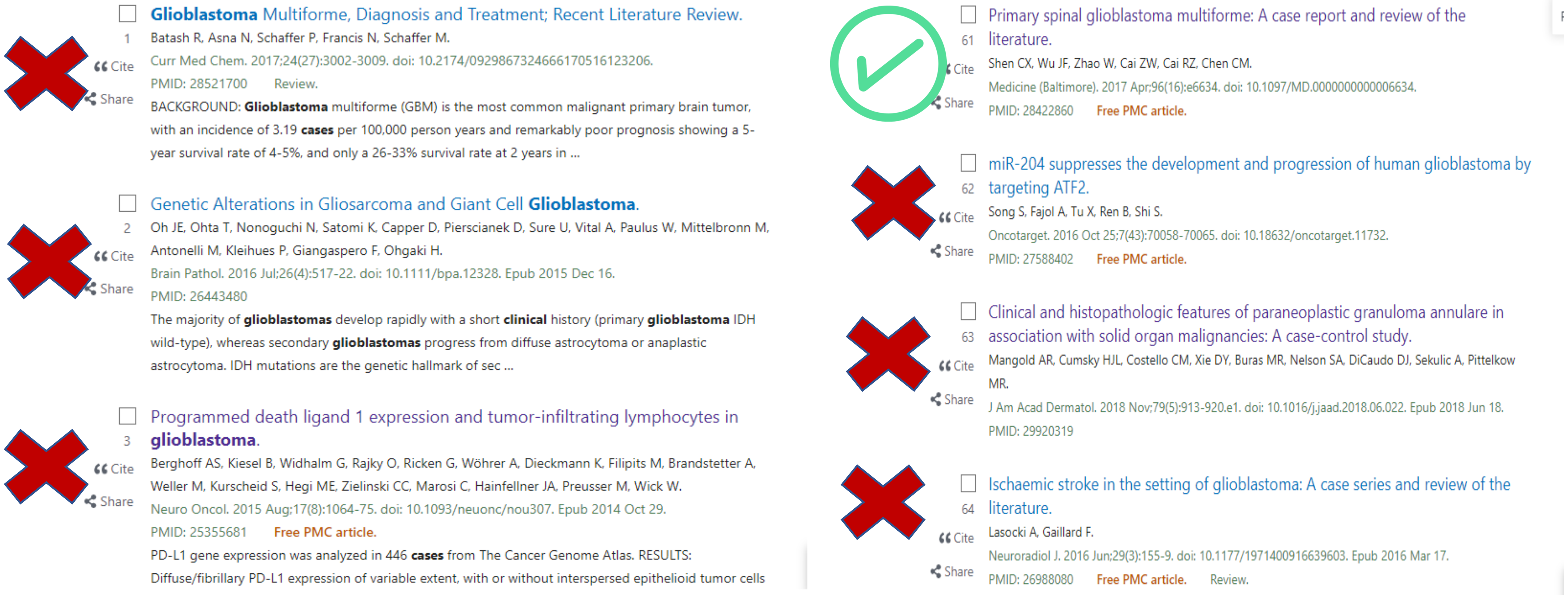

1 **Glioblastoma** Multiforme, Diagnosis and Treatment; Recent Literature Review.
Batash R, Asna N, Schaffer P, Francis N, Schaffer M.
Curr Med Chem. 2017;24(27):3002-3009. doi: 10.2174/0929867324666170516123206.
PMID: 28521700 Review.
BACKGROUND: **Glioblastoma** multiforme (GBM) is the most common malignant primary brain tumor, with an incidence of 3.19 **cases** per 100,000 person years and remarkably poor prognosis showing a 5-year survival rate of 4-5%, and only a 26-33% survival rate at 2 years in ...
Cite Share

2 Genetic Alterations in Gliosarcoma and Giant Cell **Glioblastoma**.
Oh JE, Ohta T, Nonoguchi N, Satomi K, Capper D, Pierscianek D, Sure U, Vital A, Paulus W, Mittelbronn M, Antonelli M, Kleihues P, Giangaspero F, Ohgaki H.
Brain Pathol. 2016 Jul;26(4):517-22. doi: 10.1111/bpa.12328. Epub 2015 Dec 16.
PMID: 26443480
The majority of **glioblastomas** develop rapidly with a short **clinical** history (primary **glioblastoma** IDH wild-type), whereas secondary **glioblastomas** progress from diffuse astrocytoma or anaplastic astrocytoma. IDH mutations are the genetic hallmark of sec ...
Cite Share

3 Programmed death ligand 1 expression and tumor-infiltrating lymphocytes in **glioblastoma**.
Berghoff AS, Kiesel B, Widhalm G, Rajky O, Ricken G, Wöhrer A, Dieckmann K, Filipits M, Brandstetter A, Weller M, Kurscheid S, Hegi ME, Zielinski CC, Marosi C, Hainfellner JA, Preusser M, Wick W.
Neuro Oncol. 2015 Aug;17(8):1064-75. doi: 10.1093/neuonc/nou307. Epub 2014 Oct 29.
PMID: 25355681 Free PMC article.
PD-L1 gene expression was analyzed in 446 **cases** from The Cancer Genome Atlas. RESULTS: Diffuse/fibrillary PD-L1 expression of variable extent, with or without interspersed epithelioid tumor cells
Cite Share

61 Primary spinal glioblastoma multiforme: A case report and review of the literature.
Shen CX, Wu JF, Zhao W, Cai ZW, Cai RZ, Chen CM.
Medicine (Baltimore). 2017 Apr;96(16):e6634. doi: 10.1097/MD.0000000000006634.
PMID: 28422860 Free PMC article.
Cite Share

62 miR-204 suppresses the development and progression of human glioblastoma by targeting ATF2.
Song S, Fajol A, Tu X, Ren B, Shi S.
Oncotarget. 2016 Oct 25;7(43):70058-70065. doi: 10.18632/oncotarget.11732.
PMID: 27588402 Free PMC article.
Cite Share

63 Clinical and histopathologic features of paraneoplastic granuloma annulare in association with solid organ malignancies: A case-control study.
Mangold AR, Cumsky HJL, Costello CM, Xie DY, Buras MR, Nelson SA, DiCaudo DJ, Sekulic A, Pittelkow MR.
J Am Acad Dermatol. 2018 Nov;79(5):913-920.e1. doi: 10.1016/j.jaad.2018.06.022. Epub 2018 Jun 18.
PMID: 29920319
Cite Share

64 Ischaemic stroke in the setting of glioblastoma: A case series and review of the literature.
Lasocki A, Gaillard F.
Neuroradiol J. 2016 Jun;29(3):155-9. doi: 10.1177/1971400916639603. Epub 2016 Mar 17.
PMID: 26988080 Free PMC article. Review.
Cite Share

### 3.-Eligibility.

- In total, we evaluated 108 full-text articles for eligibility. Then, we excluded 48 clinical cases articles that did not meet selection criteria such as diagnosis, Clinical follow-up of the case for at least 1 year or failing until his exitus, symptoms at the time of entering the hospital, age, sex, type of surgery or resection, treatment or therapy, regression of the tumor, molecular markers used for the identification of the GBM and life expectancy.

**Studies exclusion**

1. **Studies didn't provide the etiological information of patients or trial studies [1-6].**
2. **Studies didn't provide clinical follow-up for at least 1 year of the patient or does not comply with the required information [7-29].**
3. **Studies using cell, tumors or animal lines to test pharmacological therapies[30-37]**
4. **Studies that are clinical protocols or guidelines for the treatment of the disease[38-45].**
5. **Reviews[46-48].**

**Reference**


1. Qin JJ, Liu ZX, Wang JM, et al. Prognostic factors influencing clinical outcomes of malignant glioblastoma multiforme: clinical, immunophenotypic, and fluorescence in situ hybridization findings for 1p19q in 816 chinese cases. *Asian Pac J Cancer Prev*. 2015;16(3):971-977.

2. Sippl C, Ketter R, Braun L, et al. miRNA-26a expression influences the therapy response to carmustine wafer implantation in patients with glioblastoma multiforme. *Acta Neurochir (Wien)*. 2019;161(11):2299-2309

3. Utsuki S, Oka H, Miyajima Y, Kijima C, Yasui Y, Fujii K. Adult cerebellar glioblastoma cases have different characteristics from supratentorial glioblastoma. *Brain Tumor Pathol*. 2012;29(2):87-95.

4. Straube C, Scherb H, Gempt J, et al. Adjuvant stereotactic fractionated radiotherapy to the resection cavity in recurrent glioblastoma - the GlioCave study (NOA 17 - ARO 2016/3 - DKTK ROG trial). *BMC Cancer*. 2018;18(1):15. Published 2018 Jan 3.

5. Pompili A, Telera S, Villani V, Pace A. Home palliative care and end of life issues in glioblastoma multiforme: results and comments from a homogeneous cohort of patients. *Neurosurg Focus*. 2014;37(6):E5.

### Studies Excluded


1. Qin JJ, Liu ZX, Wang JM, et al. Prognostic factors influencing clinical outcomes of malignant glioblastoma multiforme: clinical, immunophenotypic, and fluorescence in situ hybridization findings for 1p19q in 816 chinese cases. *Asian Pac J Cancer Prev*. 2015;16(3):971-977.
2. Sippl C, Ketter R, Braun L, et al. miRNA-26a expression influences the therapy response to carmustine wafer implantation in patients with glioblastoma multiforme. *Acta Neurochir (Wien)*. 2019;161(11):2299-2309
3. Utsuki S, Oka H, Miyajima Y, Kijima C, Yasui Y, Fujii K. Adult cerebellar glioblastoma cases have different characteristics from supratentorial glioblastoma. *Brain Tumor Pathol*. 2012;29(2):87-95.
4. Straube C, Scherb H, Gempt J, et al. Adjuvant stereotactic fractionated radiotherapy to the resection cavity in recurrent glioblastoma - the GlioCave study (NOA 17 - ARO 2016/3 - DKTK ROG trial). *BMC Cancer*. 2018;18(1):15. Published 2018 Jan 3.
5. Pompili A, Telera S, Villani V, Pace A. Home palliative care and end of life issues in glioblastoma multiforme: results and comments from a homogeneous cohort of patients. *Neurosurg Focus*. 2014;37(6):E5.
6. Liu XR, Hsiao TY, Li YQ, et al. Neurosurgical brain tumor detection based on intraoperative optical intrinsic signal imaging technique: A case report of glioblastoma. *J Biophotonics*. 2020;13(1):e201900200.
7. Mahvash M, Hugo HH, Maslehaty H, Mehdorn HM, Stark AM. Glioblastoma multiforme in children: report of 13 cases and review of the literature. *Pediatr Neurol*. 2011;45(3):178-180.
8. O'Halloran PJ, Farrell M, Caird J, Capra M, O'Brien D. Paediatric spinal glioblastoma: case report and review of therapeutic strategies. *Childs Nerv Syst*. 2013;29(3):367-374.
9. Van Ginderachter L, Cox T, Drijkoningen R, et al. Non-hodgkin lymphoma after treatment with extended dosing temozolomide and radiotherapy for a glioblastoma: a case report. *Case Rep Oncol*. 2013;6(1):45-49.
10. Rühle PF, Goerig N, Wunderlich R, et al. Modulations in the Peripheral Immune System of Glioblastoma Patient Is Connected to Therapy and Tumor Progression-A Case Report from the IMMO-GLIO-01 Trial. *Front Neurol*. 2017;8:296.
11. Baillie N, Carr AC, Peng S. The Use of Intravenous Vitamin C as a Supportive Therapy for a Patient with Glioblastoma Multiforme. *Antioxidants (Basel)*. 2018;7(9):115.
12. Caro-Osorio E, Herrera-Castro JC, Barbosa-Quintana A, Benvenutti-Regato M. Primary Spinal Cord Small-Cell Glioblastoma: Case Report and Literature Review. *World Neurosurg*. 2018;118:69-70.
13. Tejada S, Díez-Valle R, Domínguez PD, et al. DNX-2401, an Oncolytic Virus, for the Treatment of Newly Diagnosed Diffuse Intrinsic Pontine Gliomas: A Case Report. *Front Oncol*. 2018;8:61.
14. Bärtschi P, Luna E, González-López P, et al. A Very Rare Case of Right Insular Lobe Langerhans Cell Histiocytosis ($CD1a^{+}$) Mimicking Glioblastoma Multiforme in a Young Adult. *World Neurosurg*. 2019;121:4-11.
15. DeDios-Stern S, Durkin NM, Soble JR. Case of right hemispatial neglect and transcortical sensory aphasia following left occipitotemporoparietal glioblastoma resection [published online ahead of print, 2019 Apr 15]. *Appl Neuropsychol Adult*. 2019;1-7.
16. Piper K, Foster H, Gabel B, Nabors B, Cobbs C. Glioblastoma Mimicking Viral Encephalitis Responds to Acyclovir: A Case Series and Literature Review. *Front Oncol*. 2019;9:8. Published 2019 Jan 22.
17. Naydenov E, Bussarsky V, Minkin K, Bussarsky A, Nachev S, Traykov L. Normal pressure hydrocephalus as an unusual presentation of supratentorial extraventricular space-occupying processes: report on two cases. *Case Rep Oncol*. 2012;5(1):143-147.
18. Arcos, A., Romero, L., Serramito, R., Santín, J. M., Prieto, A., Gelabert, M., & Arráez, M. Á. (2012). *Multicentric glioblastoma multiforme. Report of 3 cases, clinical and pathological study and literature review. Neurocirugía, 23(5), 211*
19. Deng S, Liu L, Wang D, Tong D, Zhao G. Small Cell Glioblastoma of the Sella Turcica Region: Case Report and Review of the Literature. *World Neurosurg*. 2018;110:174-179. doi:10.1016/j.wneu.2017.11.038
20. Eva L, Dobrovăţ BI, Haba D, et al. A novel combination of double primary malignancies: penile carcinoma and glioblastoma. A series of two cases. *Rom J Morphol Embryol*. 2018;59(4):1067-1074.
21. MacMahon P, Labak CM, Martin-Bach SE, Issawi A, Velpula K, Tsung AJ. Glioblastoma formation in a recurrent intracranial epidermoid cyst: a case report. *CNS Oncol*. 2018;7(4):CNS25.
22. Hammas N, Senhaji N, Alaoui Lamrani MY, et al. Astroblastoma - a rare and challenging tumor: a case report and review of the literature. *J Med Case Rep*. 2018;12(1):102. Published 2018 Apr 21.
23. Chehimi M, Boone M, Chivot C, et al. Intra-Arterial Delivery of Idarubicin in Two Patients with Glioblastoma. *Case Rep Oncol*. 2016;9(2):499-505. Published 2016 Aug 30.
24. Munhoz RR, Pereira Picarelli AA, Troques Mitteldorf CA, Feher O. Aspergillosis in a patient receiving temozolomide for the treatment of glioblastoma. *Case Rep Oncol*. 2013;6(2):410-415. Published 2013 Aug 8.
25. Konar SK, Shukla D, Nandeesh BN, Prabhuraj AR, Devi BI. Surgical Management and Outcome of a Bilateral Thalamic Pilocytic Astrocytoma: Case Report and Review of the Literature. *Pediatr Neurosurg*. 2019;54(2):139-142.
26. Thumma SR, Elaimy AL, Daines N, et al. Long-term survival after gamma knife radiosurgery in a case of recurrent glioblastoma multiforme: a case report and review of the literature. *Case Rep Med*. 2012;2012:545492.
27. Lin CY, Huang HM. Unilateral malignant optic glioma following glioblastoma multiforme in the young: a case report and literature review. *BMC Ophthalmol*. 2017;17(1):21. Published 2017 Mar 11.
28. Hanaei S, Habibi Z, Nejat F, Sayarifard F, Vasei M. Pediatric glioblastoma multiforme in association with Turner's syndrome: a case report. *Pediatr Neurosurg*. 2015;50(1):38-41.
29. Van Ginderachter L, Cox T, Drijkoningen R, et al. Non-hodgkin lymphoma after treatment with extended dosing temozolomide and radiotherapy for a glioblastoma: a case report. *Case Rep Oncol*. 2013;6(1):45-49.
30. Lu VM, Kerezoudis P, Brown DA, Burns TC, Quinones-Hinojosa A, Chaichana KL. Hypofractionated versus standard radiation therapy in combination with temozolomide for glioblastoma in the elderly: a meta-analysis. *J Neurooncol*. 2019;143(2):177-185.
31. Berghoff AS, Kiesel B, Widhalm G, et al. Programmed death ligand 1 expression and tumor-infiltrating lymphocytes in glioblastoma. *Neuro Oncol*. 2015;17(8):1064-1075. doi:10.1093/neuonc/nou307
32. Qiang Z, Jun-Jie L, Hai W, et al. TPD52L2 impacts proliferation, invasiveness and apoptosis of glioblastoma cells via modulation of wnt/β-catenin/snail signaling. *Carcinogenesis*. 2018;39(2):214-224.
33. Miyai M, Tomita H, Soeda A, Yano H, Iwama T, Hara A. Current trends in mouse models of glioblastoma. *J Neurooncol*. 2017;135(3):423-432.
34. Urdiciain A, Erausquin E, Meléndez B, Rey JA, Idoate MA, Castresana JS. Tubastatin A, an inhibitor of HDAC6, enhances temozolomide-induced apoptosis and reverses the malignant phenotype of glioblastoma cells. *Int J Oncol*. 2019;54(5):1797-1808.
35. Takahashi Y, Makino K, Nakamura H, et al. Clinical characteristics and pathogenesis of cerebellar glioblastoma. *Mol Med Rep*. 2014;10(5):2383-2388.
36. Korshunov A, Casalini B, Chavez L, et al. Integrated molecular characterization of IDH-mutant glioblastomas. *Neuropathol Appl Neurobiol*. 2019;45(2):108-118.
37. Rao S, Kanuri NN, Nimbalkar V, Arivazhagan A, Santosh V. High frequency of H3K27M immunopositivity in adult thalamic glioblastoma. *Neuropathology*. 2019;39(2):78-84.
38. D'Alessandris QG, Biffoni M, Martini M, et al. The clinical value of patient-derived glioblastoma tumorspheres in predicting treatment response. *Neuro Oncol*. 2017;19(8):1097-1108.
39. Karlowee V, Amatya VJ, Takayasu T, et al. Immunostaining of Increased Expression of Enhancer of Zeste Homolog 2 (EZH2) in Diffuse Midline Glioma H3K27M-Mutant Patients with Poor Survival. *Pathobiology*. 2019;86(2-3):152-161.
40. Smith DR, Wu CC, Saadatmand HJ, et al. Clinical and molecular characteristics of gliosarcoma and modern prognostic significance relative to conventional glioblastoma. *J Neurooncol*. 2018;137(2):303-311.
41. Mansouri A, Hachem LD, Mansouri S, et al. MGMT promoter methylation status testing to guide therapy for glioblastoma: refining the approach based on emerging evidence and current challenges. *Neuro Oncol*. 2019;21(2):167-178.
42. Brandes AA, Franceschi E, Paccapelo A, et al. Role of *MGMT* Methylation Status at Time of Diagnosis and Recurrence for Patients with Glioblastoma: Clinical Implications. *Oncologist*. 2017;22(4):432-437.
43. Barritault M, Meyronet D, Ducray F. Molecular classification of adult gliomas: recent advances and future perspectives. *Curr Opin Oncol*. 2018;30(6):375-382.
44. Esquenazi Y, Moussazadeh N, Link TW, et al. Thalamic Glioblastoma: Clinical Presentation, Management Strategies, and Outcomes. *Neurosurgery*. 2018;83(1):76-85.
45. Vogrig A, Joubert B, Ducray F, et al. Glioblastoma as differential diagnosis of autoimmune encephalitis. *J Neurol*
46. Batash R, Asna N, Schaffer P, Francis N, Schaffer M. Glioblastoma Multiforme, Diagnosis and Treatment; Recent Literature Review. *Curr Med Chem*. 2017;24(27):3002-3009.
47. Eder K, Kalman B. Molecular heterogeneity of glioblastoma and its clinical relevance. *Pathol Oncol Res*. 2014;20(4):777-787. doi:10.1007/s12253-014-9833-3
48. Goryaynov SA, Potapov AA, Ignatenko MA, et al. Glioblastoma metastases: a literature review and a description of six clinical observations. *Zh Vopr Neirokhir Im N N Burdenko*. 2015;79(2):33-43. doi:10.17116/neiro201579233-43

### 4.-Included

- In total, we included 60 publications. The studies were published between 2005 and 2020 and reported on 73 patients. The procedure of publication retrieval and in- and exclusion of cases is displayed in a PRISMA (Preferred Reporting Items for Systematic Reviews and Meta-Analyzes)

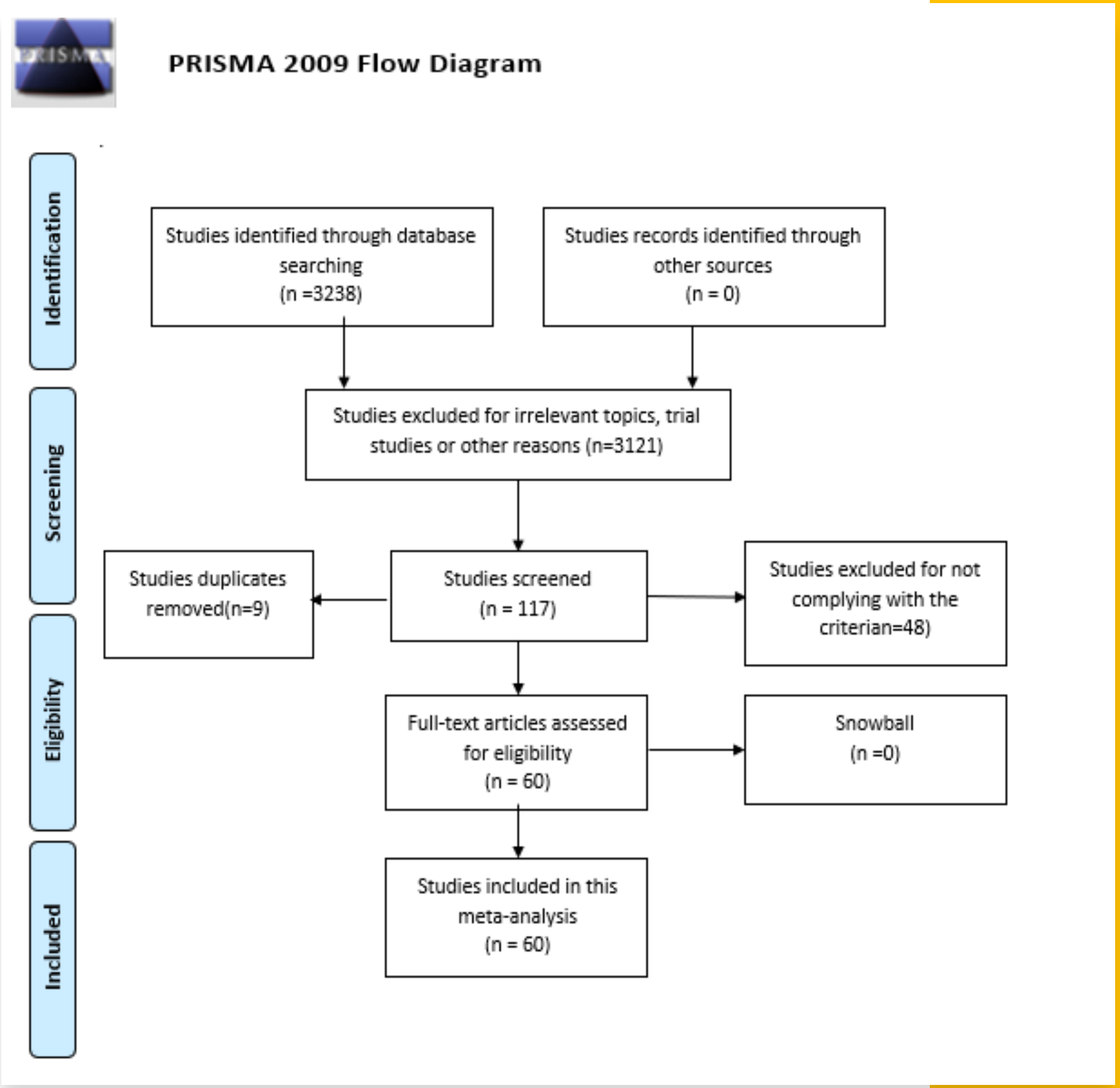

### 5. Data extraction

**5.1 From the eligible articles, the following variables were recorded on a standard data extraction form:**

1. Reference of the clinical report year of publication
2. Database where the clinical report was found
3. Sex of the patient of the clinical case reported
4. Age of the patient of the clinical case reported.
5. Symptoms described during clinical routine.
6. GBM tumor according to ontogenesis subtypes.
7. Surgical procedures during patient treatment.
8. Medical and drugs administrated for treatment.
9. Reccurence of tumor after surgical procedures
10. Gene expression and mutations measured for diagnosis
11.Survival time of patients after surgery and teraphy treatment.

### 5.1 Data directly extracted from literatures

2 ➡

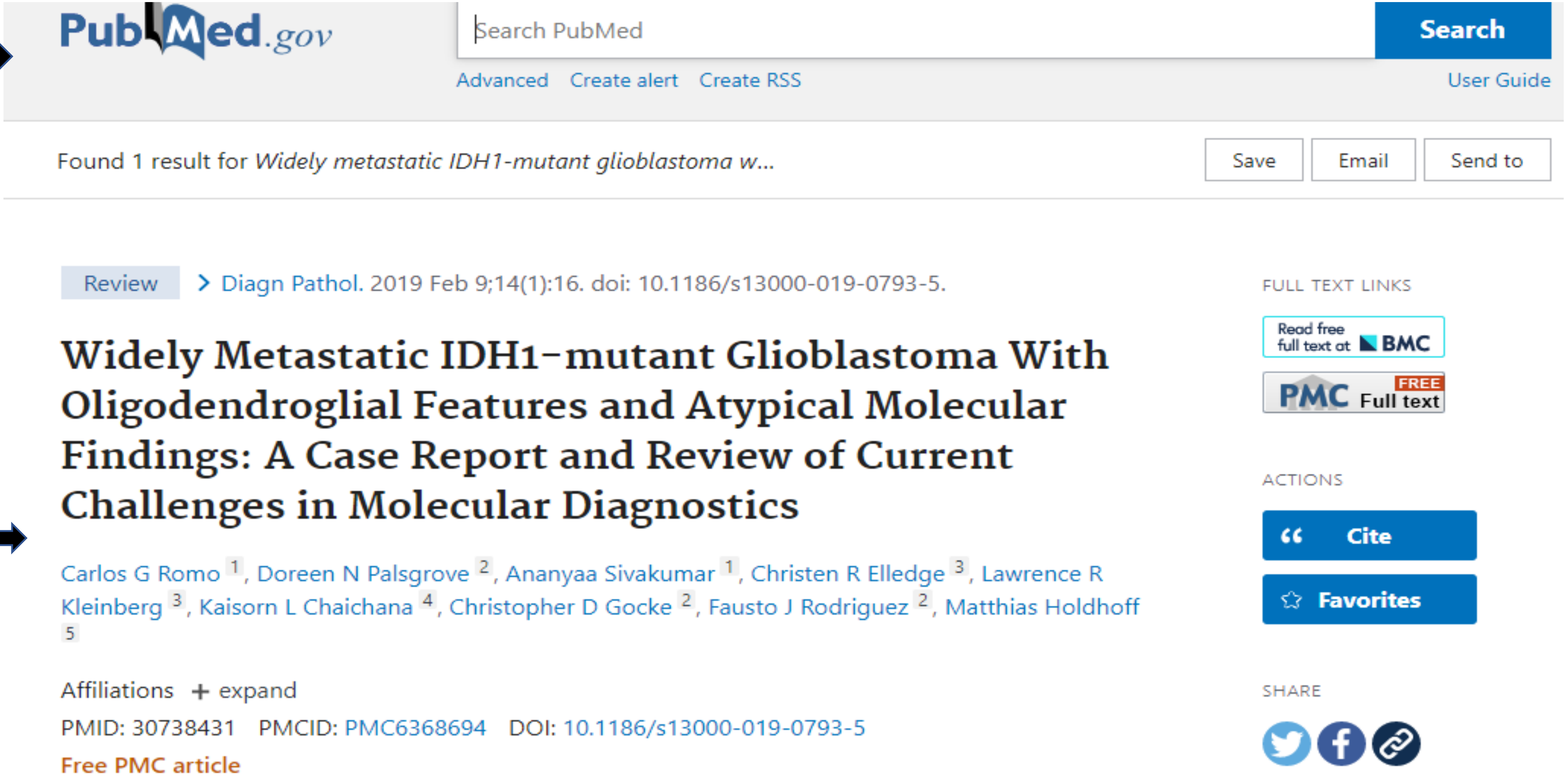

PubMed.gov

Search PubMed | Search

Advanced   Create alert   Create RSS   User Guide

Found 1 result for *Widely metastatic IDH1-mutant glioblastoma w...*   Save   Email   Send to

Review  > Diagn Pathol. 2019 Feb 9;14(1):16. doi: 10.1186/s13000-019-0793-5.

**Widely Metastatic IDH1-mutant Glioblastoma With Oligodendroglial Features and Atypical Molecular Findings: A Case Report and Review of Current Challenges in Molecular Diagnostics**

Carlos G Romo [1], Doreen N Palsgrove [2], Ananyaa Sivakumar [1], Christen R Elledge [3], Lawrence R Kleinberg [3], Kaisorn L Chaichana [4], Christopher D Gocke [2], Fausto J Rodriguez [2], Matthias Holdhoff [5]

Affiliations + expand

PMID: 30738431   PMCID: PMC6368694   DOI: 10.1186/s13000-019-0793-5

Free PMC article

FULL TEXT LINKS

Read free full text at BMC

PMC FREE Full text

ACTIONS

Cite

Favorites

SHARE

1 ➡

### 5.1 Data directly extracted from literatures (Example in PubMed Database )

#### Case presentation

5

The patient was a 28-year-old man who presented with a 2-week history of subtle personality changes, expressive aphasia, and severe headaches with nausea and vomiting. Imaging revealed a large mass in the left frontal lobe with significant surrounding vasogenic edema (Fig [illegible] 7 He underwent gross total resection and pathology was consistent with a high grade neuroepithelial tumor with brisk mitotic activity and necrosis (Fig. 2, Additional file 1 : Figures S1 a[illegible] 10 ). The tumor was found to be *IDH1* and *TP53* mutant by amplicon-based next generation sequencing (NGS) targeting the hotspot regions of 27 cancer associated genes (not including *ATRX*), and had monosomy 1 and 19q loss by single nucleotide polymorphism (SNP) array (Figs. 2 and 3). A diagnosis of anaplastic oligodendroglioma was favored based on histologic appearance of the majority of the tissue with features suggestive of an oligodendroglioma in addition to preserved ATRX immunoreactivity, and atypical alterations in chromosomes 1 and 19. Treatment options were discussed including participation in the CODEL trial (NCT00887146), but ultimately started standard of care treatment with radiation therapy with plan for adjuvant procarbazine, lomustine, and vincristine (PCV) chemotherapy following radiation. Unfortunately, after resection and prior to starting radiation he developed somnolence, an intense headache, and diplopia. MRI of the brain revealed evidence of tumor regrowth in the original location and signs of impending herniation. He was started on dexamethasone and underwent emergent surgical decompression with tumor debulking. Pathology was again consistent with a high-grade *IDH1*-mutant glioma. His symptoms improved and he received intensity-modulated radiation therapy (IMRT) with a cumulative dose of 5940 cGy over 33 fractions. MGMT promoter methylation was tested upon recurrence and was found to be present.

In addition to tumor lysis, the patient rapidly became thrombocytopenic (179 K/cu mm on admission to a nadir of 35 K/cu mm by day seven) and anemic (14.5 g/dL to a nadir of 5.8 g/dL), likely due to bone marrow infiltration of his cancer. Different chemotherapy regimens were considered and the choice was for temozolomide, as it is typically effective in diffuse gliomas and is able to cross the blood-brain barrier with even higher concentrations systemically. Five doses were planned (150 mg/m$^2$) but the patient was only able to receive two administrations.

Unfortunately, the patient's condition continued to deteriorate. He developed worsening renal failure, hypotension, and acute respiratory failure. He was intubated and mechanically ventilated, requiring vasopressor support. The patient developed cardiac arrest, was

11 resuscitated and eventually transitioned to comfort measures. He was terminally extubated and passed away. An autopsy was declined.

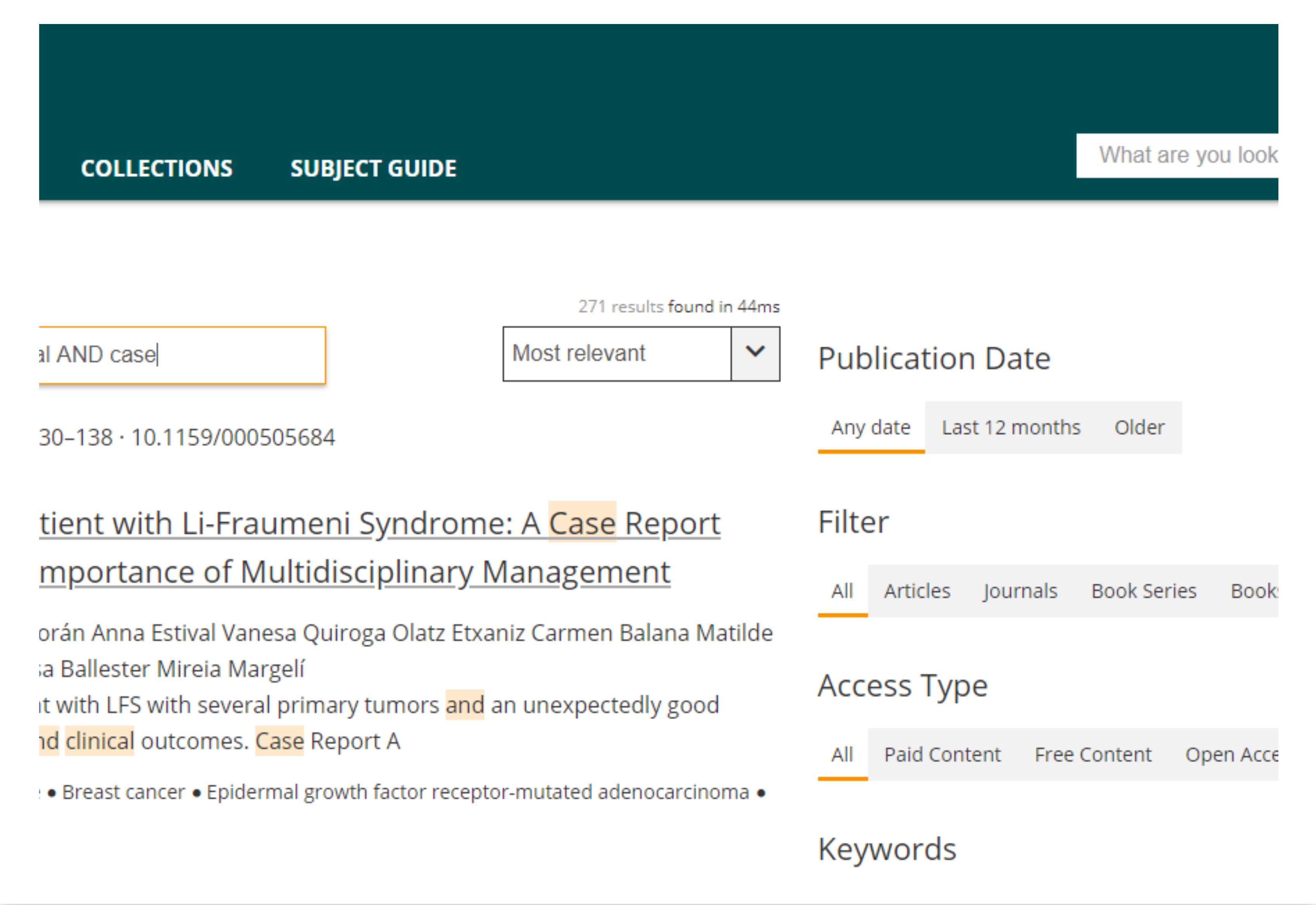


#### 5.1 Data directly extracted from literatures(Example in Karger)

DOWNLOAD FULLTEXT PDF

Case Reports in Oncology

Open access

Case Report

2 ➡ 

### Extracranial Metastases of a Cerebral Glioblastoma: A Case Report and Review of the Literature

1 ➡

Rosen J.[a] · Blau T.[b] · Grau S.J.[c] · Barbe M.T.[a] · Fink G.R.[a,d] · Galldiks N.[a,d,e]
Author affiliations

Corresponding Author

Keywords: Glioma · Extraneural metastasis · Radiotherapy · Chemotherapy · Tumor resection

Case Rep Oncol 2018;11:591–600
https://doi.org/10.1159/000492111

ABSTRACT FULLTEXT PDF REFERENCES EXTRAS : 4

Article Tools

Get Permission
PubMed ID
Citation Download
Web of Science Citations: 10
Add to my Reading List
4

Article Details

2018, Vol.11, No. 2
May – August
Open access

PREV Article NEXT

Recommend This

Case Reports in Oncology

Case Rep Oncol 2018;11:591–600

DOI: 10.1159/000492111

© 2018 The Author(s). Published by S. Karger AG, Basel
www.karger.com/cro



**Case Presentation**

6

3,4 A 48-year-old female patient with no significant past medical history presented with progressive cognitive deficits, deterioration of speech, and a new right-sided hemiparesis. Furthermore, a newly diagnosed right-sided hemiparesis led to a cerebral MRI and revealed a left temporal lesion suspicious of a GBM ([illegible]). 7 After partial resection, the symptoms improved, and neuropathological [illegible] 10 confirmed a GBM (MGMT promoter not methylated, IDH-1/-2 wild type). Aft[illegible] 8 s, radiotherapy (total dose, 60 Gy) with concomitant temozolomide chemotherapy (daily dose of 150 mg over 6 weeks) was initiated. Due to unwanted and intolerable side effects (i.e., loss of appetite, nausea, vomiting), adjuvant temozolomide chemotherapy was paused after the first cycle.

5

Two months later, the patient presented with recurrent disturbance of speech and 9 memory deficits. Follow-up MRI revealed local tumor progression, and a second resection was performed. Neuropathological findings were consistent with a GBM relapse. Consequently, adjuvant chemotherapy with temozolomide was re-initiated, which was now tolerated and followed by an improvement of speech and cognition. Five months after the initial diagnosis of the GBM, the patient complained of a persistent local pain in the left maxilla. The assumption of an underlying dental pathology led to a dental extraction, which, however, did not remove the pain. Subsequent MR imaging rather showed a tumor progression through the skull base into the pterygopalatine fossa with infiltration of the soft tissue (Fig. 1b). Additionally, a lymph node metastasis of the neck was detected, most probably due to a GBM tumor cell infiltration of local lymphatic vessels (Fig. 1c).

Consequently, a second palliative radiotherapy with a cumulative dose of 52 Gy of the neck region followed by adjuvant chemotherapy with lomustine was initiated. Despite these efforts, the patient suffered from progressive speech disturbances and nausea, which could be explained by an extensive progression of the tumor lesion inside and outside the cranium (Fig. 1d). Antiangiogenic therapy with bevacizumab was initiated.

Only 3 weeks after the initiation of bevacizumab treatment, the patient suffered from a severe intestinal hemorrhage. Despite a CT scan of the thorax and abdomen, colono-, and gastroscopy, no source of the bleeding could be detected. Fortunately, the bleeding ceased spontaneously.

Unexpectedly, however, the CT scan detected multiple lesions in the lung, pleura, liver, mesentery as well as osteolyses of the pelvis, the seventh rib on the left, and the second lumbar vertebral body, all suspicious of being metastases (Fig. 2). Moreover, a suspect subcutaneous

lesion located at the right upper arm was palpable. The pathological examination of another subcutaneous lesion located below the left costal arch demonstrated the same histological characteristics as the intracranial GBM (Fig. 3). Subsequently, the patient was admitted to palliative care and died due to intracranial progression of the tumor 13 months after initial diagnosis.

11

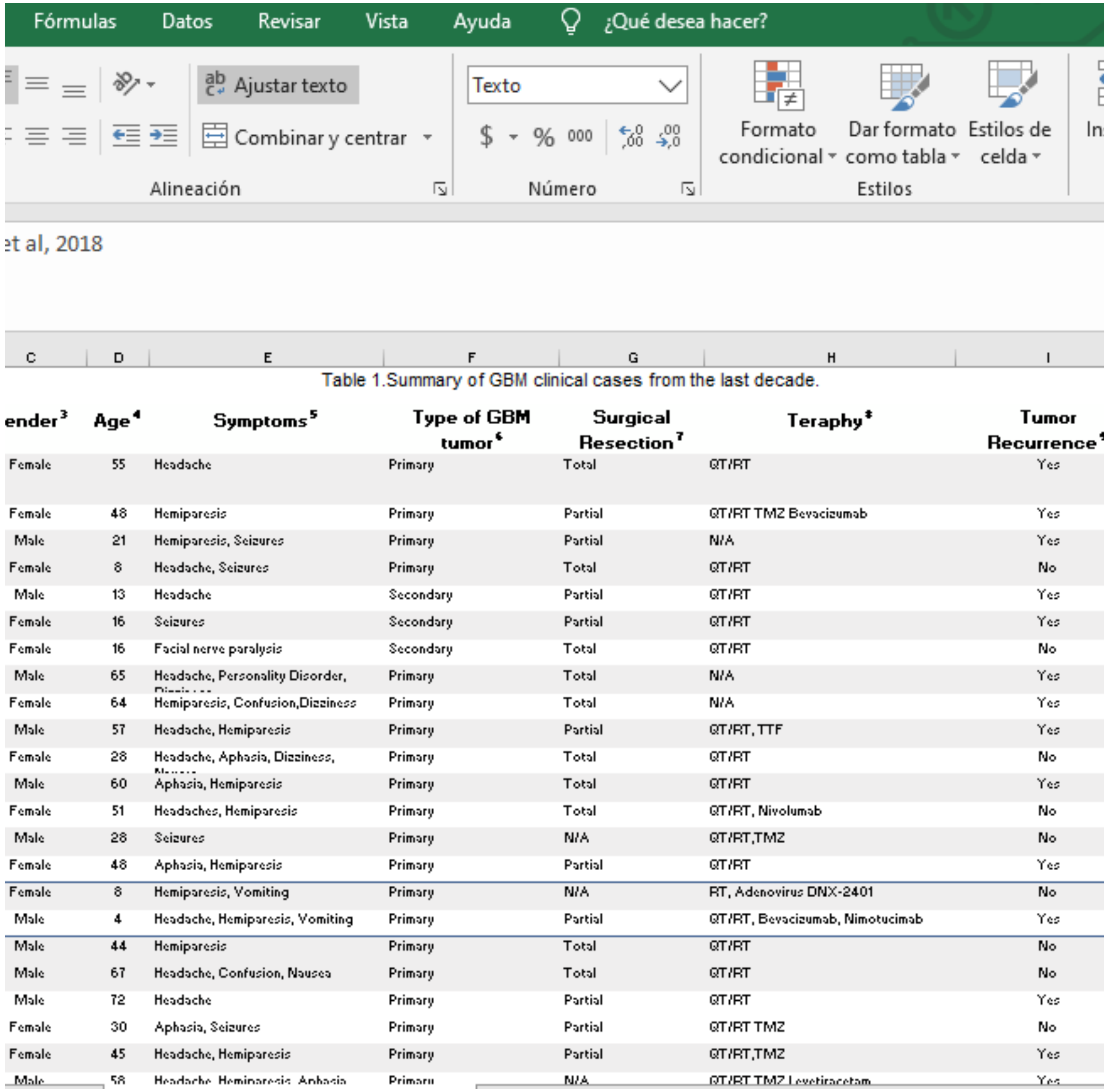


Table 1.Summary of GBM clinical cases from the last decade.

| ender[3] | Age[4] | Symptoms[5] | Type of GBM tumor[6] | Surgical Resection[7] | Teraphy[8] | Tumor Recurrence[illegible] |
|---|---|---|---|---|---|---|
| Female | 55 | Headache | Primary | Total | QT/RT | Yes |
| Female | 48 | Hemiparesis | Primary | Partial | QT/RT TMZ Bevacizumab | Yes |
| Male | 21 | Hemiparesis, Seizures | Primary | Partial | N/A | Yes |
| Female | 8 | Headache, Seizures | Primary | Total | QT/RT | No |
| Male | 13 | Headache | Secondary | Partial | QT/RT | Yes |
| Female | 16 | Seizures | Secondary | Partial | QT/RT | Yes |
| Female | 16 | Facial nerve paralysis | Secondary | Total | QT/RT | No |
| Male | 65 | Headache, Personality Disorder, [illegible] | Primary | Total | N/A | Yes |
| Female | 64 | Hemiparesis, Confusion,Dizziness | Primary | Total | N/A | Yes |
| Male | 57 | Headache, Hemiparesis | Primary | Partial | QT/RT, TTF | Yes |
| Female | 28 | Headache, Aphasia, Dizziness, [illegible] | Primary | Total | QT/RT | No |
| Male | 60 | Aphasia, Hemiparesis | Primary | Total | QT/RT | Yes |
| Female | 51 | Headaches, Hemiparesis | Primary | Total | QT/RT, Nivolumab | No |
| Male | 28 | Seizures | Primary | N/A | QT/RT,TMZ | No |
| Female | 48 | Aphasia, Hemiparesis | Primary | Partial | QT/RT | Yes |
| Female | 8 | Hemiparesis, Vomiting | Primary | N/A | RT, Adenovirus DNX-2401 | No |
| Male | 4 | Headache, Hemiparesis, Vomiting | Primary | Partial | QT/RT, Bevacizumab, Nimotucimab | Yes |
| Male | 44 | Hemiparesis | Primary | Total | QT/RT | No |
| Male | 67 | Headache, Confusion, Nausea | Primary | Total | QT/RT | No |
| Male | 72 | Headache | Primary | Partial | QT/RT | Yes |
| Female | 30 | Aphasia, Seizures | Primary | Partial | QT/RT TMZ | No |
| Female | 45 | Headache, Hemiparesis | Primary | Partial | QT/RT,TMZ | Yes |
| Male | 58 | Headache, Hemiparesis, Aphasia | Primary | N/A | QT/RT TMZ Levetiracetam | Yes |

### Meta-analysis for clinicopathological parameters

### Studies included in the meta analysis


1. Anghileri E, Castiglione M, Nunziata R, et al. Extraneural metastases in glioblastoma patients: two cases with YKL-40-positive glioblastomas and a meta-analysis of the literature [published correction appears in Neurosurg Rev. 2015 Oct;38(4):773. Elena, Anghileri [corrected to Anghileri, Elena]; Melina, Castiglione [corrected to Castiglione, Melina]; Raffaele, Nunziata [corrected to Nunziata, Raffaele]; Carlo, Boffano [corrected to Boffano, Carlo]; Vittoria, Nazzi [corrected to Nazzi, Vittoria]; Fr]. *Neurosurg Rev*. 2016;39(1):37-46.
2. Awadalla AS, Al Essa AM, Al Ahmadi HH, et al. Gliosarcoma case report and review of the literature. *Pan Afr Med J*. 2020;35:26. Published 2020 Feb 3.
3. Bärtschi, P., Luna, E., González-López, P., Abarca, J., Herrero, J., Costa, E., ... & Moreno, P. (2019). A Very Rare Case of Right Insular Lobe Langerhans Cell Histiocytosis (CD1a+) Mimicking Glioblastoma Multiforme in a Young Adult. *World neurosurgery*, *121*, 4-11.
4. Chanchotisatien A, Xiong J, Yu J, Chu S. Exophytic Primary Intramedullary Spinal Cord Glioblastoma: Case Report and Critical Review of Literature. *World Neurosurg*. 2019;122:573-576.
5. Chen, F., Li, Z., Weng, C., Li, P., Tu, L., Chen, L., ... & Li, L. (2017). Glioblastoma pontino exofítico multifocal progresivo: reporte de un caso con revisión de la literatura. *Revista china de cáncer* , *36* (1), 34.
6. Comito, R. R., Badu, L. A., & Forcello, N. (2019). Nivolumab-induced aplastic anemia: a case report and literature review. *Journal of Oncology Pharmacy Practice*, *25*(1), 221-225.
7. Cuoco JA, Klein BJ, Busch CM, Guilliams EL, Olasunkanmi AL, Entwistle JJ. Corticosteroid-Induced Regression of Glioblastoma: A Radiographic Conundrum. *Front Oncol*. 2019;9:1288. Published 2019 Nov 22.
8. Dormegny, L., Chibbaro, S., Ganau, M., Santin, M. D. N., Kremer, L., & Proust, F. (2018). Biopsying a spinal cord lesion: a diagnostic dilemma. Case report and review of literature. *Neurochirurgie*, *64*(6), 425-430.
9. Efferth T, Schöttler U, Krishna S, Schmiedek P, Wenz F, Giordano FA. Hepatotoxicity by combination treatment of temozolomide, artesunate and Chinese herbs in a glioblastoma multiforme patient: case report review of the literature. *Arch Toxicol*. 2017;91(4):1833-1846.
10. Elzinga, G., & Wong, E. T. (2014). Resolution of cystic enhancement to add-on tumor treating electric fields for recurrent glioblastoma after incomplete response to bevacizumab. Case reports in neurology, 6(1), 109-115.
11. Finneran, M. M., Georges, J., Kakareka, M., Moncman, R., Enriquez, M., Siegal, T., ... & Barrese, J. (2019). Epithelioid glioblastoma presenting as aphasia in a young adult with ovarian cancer: A case report. *Clinical case reports*, *7*(4), 821.
12. Gandhi, P., Khare, R., Garg, N., & Sorte, S. (2017). Immunophenotypic signature of primary glioblastoma multiforme: A case of extended progression free survival. *World journal of clinical cases*, *5*(6), 247.
13. Gestrich C, Cowden D, Harbhajanka A. Cytomorphology of glioblastoma metastic to a cervical lymph node diagnosed by fine needle aspiration (FNA): A case report and review of literature. *Diagn Cytopathol*. 2020;48(6):567-570.
14. Gupta, S., Guha, D., Taslimi, S., Priola, S. M., Waggass, G., Heyn, C., ... & Cook, D. J. (2020). Multi-modality imaging assisted fluorescence-guided resection of glioblastoma: Case report. *Interdisciplinary Neurosurgery*, *19*, 100593.
15. Hasan, S., Gigliotti, M. J., Deutsch, M., Reed, S. L., & Wegner, R. E. (2018). A 58-Year-Old Woman with Left-Sided Weakness and a History of a Pediatric Brain Tumor: A Case Report. *Case reports in oncology*, *11*(1), 131-137

16. He, M. X., & Wang, J. J. (2011). Rhabdoid glioblastoma: case report and literature review. *Neuropathology*, *31*(4), 421-426.

17. Homma, T., Hanashima, Y., Maebayashi, T., Nakanishi, Y., Ishige, T., Ohta, T., ... & Hao, H. (2019). Papillary glioblastoma exhibiting a neuroradiological cyst with a mural nodule: A case report. *Medicine*, *98*(2).

18. Hou, LC, Bababeygy, SR, Sarkissian, V., Fisher, PG, Vogel, H., Barnes, P. y Huhn, SL (2008). Congenital glioblastoma multiforme: case report and literature review. *Pediatric neurosurgery, 44* (4), 304-312.

19. Janik, K., Och, W., Popeda, M., Rosiak, K., Peciak, J., Rieske, P., ... y Stoczynska-Fidelus, E. (2019). Glioblastoma with BRAFV600E mutation and numerous metastatic foci: a case report. *Neuropathological* , 57 (1), 72-79.

20. Jeong, T. S., & Yee, G. T. (2014). Glioblastoma in a patient with neurofibromatosis type 1: a case report and review of the literature. *Brain tumor research and treatment*, *2*(1), 36-38.

21. Johansen MD, Rochat P, Law I, Scheie D, Poulsen HS, Muhic A. Presentation of Two Cases with Early Extracranial Metastases from Glioblastoma and Review of the Literature. *Case Rep Oncol Med*. 2016;2016:8190950.

22. Johnson, DR, Fogh, SE, Giannini, C., Kaufmann, TJ, Raghunathan, A., Theodosopoulos, PV, y Clarke, JL (2015). Case-based review: newly diagnosed glioblastoma. *Neuro-oncology practice*,*2* (3), 106-121.

23. Kajitani, T., Kanamori, M., Saito, R., Watanabe, Y., Suzuki, H., Watanabe, M., ... & Tominaga, T. (2018). Three case reports of radiation-induced glioblastoma after complete remission of acute lymphoblastic leukemia. *Brain tumor pathology*, *35*(2), 114-122.

24. Kumaria, A., Teale, A., Kulkarni, G. V., Ingale, H. A., Macarthur, D. C., & Robertson, I. J. (2018). Glioblastoma multiforme metastatic to lung in the absence of intracranial recurrence: case report. *British journal of neurosurgery*, 1-3.

25. Lakičević G, Arnautović K, Mužević D, Chesney T. Cerebellar glioblastoma multiforme presenting as hypertensive cerebellar hemorrhage: case report. *J Neurol Surg Rep*. 2014;75(1):e117-e121.

26. Lewis, G. D., Rivera, A. L., Tremont-Lukats, I. W., Ballester-Fuentes, L. Y., Zhang, Y. J., & Teh, B. S. (2017). GBM skin metastasis: a case report and review of the literature. *CNS oncology*, *6*(3), 203-209.

27. Macchi ZA, Kleinschmidt-DeMasters BK, Orjuela KD, Pastula DM, Piquet AL, Baca CB. Glioblastoma as an autoimmune limbic encephalitis mimic: A case and review of the literature [published online ahead of print, 2020 Mar 7]. *J Neuroimmunol*. 2020;342:577214.

28. Matsuda M, Onuma K, Satomi K, Nakai K, Yamamoto T, Matsumura A. Exophytic cerebellar glioblastoma in the cerebellopontine angle: case report and review of the literature. *J Neurol Surg Rep*. 2014;75(1):e67-e72.

29. McClelland III, S., Henrikson, C. A., Ciporen, J. N., Jaboin, J. J., & Mitin, T. (2018). Tumor treating fields utilization in a glioblastoma patient with a preexisting cardiac pacemaker: The first reported case. *World neurosurgery*, *119*, 58-60.

30. Narasimhaiah, D., Sridutt, B. S., Thomas, B., & Vilanilam, G. C. (2019). Glioblastoma in adults with neurofibromatosis type I: A report of two cases. *Neuropathology*, *39*(5), 368-373.

31. Naydenov, E., Bussarsky, V., Angelov, K., Penkov, M., Nachev, S., Hadjidekova, S. y Toncheva, D. (2017). Metachronic development of meningothelial meningioma, basal cell carcinoma and glioblastoma multiforme in a patient with pancreatic incidentaloma. *Case reports in oncology*, *10* (3), 1023-1028.

32. Naydenov, E., Bussarsky, V., Nachev, S., Hadjidekova, S. y Toncheva, D. (2009). Long-term survival of a patient with giant cell glioblastoma: case report and literature review. *Case reports in oncology, 2* (2), 103-110.

33. Nørøxe, D. S., Michaelsen, S. R., Broholm, H., Møller, S., Poulsen, H. S., & Lassen, U. (2019). Extracranial metastases in glioblastoma—Two case stories. *Clinical case reports*, *7*(2), 289.

34. Papaevangelou G, Chamilos C, Dana H, Sgouros S(2017): Cerebellar Glioblastoma in a Child. Pediatr Neurosurg 201752:211-213.

35. Paraskevopoulos, D., Patsalas, I., Karkavelas, G., Foroglou, N., Magras, I., & Selviaridis, P. (2011). Pilomyxoid astrocytoma of the cervical spinal cord in a child with rapid progression into glioblastoma: case report and literature review. *Child's Nervous System*, *27*(2), 313-321.

36. Petzold, J., Severus, E., Meyer, S., Bauer, M., Daubner, D., Krex, D., & Juratli, T. A. (2018). Glioblastoma multiforme presenting as postpartum depression: a case report. *Journal of medical case reports*, *12*(1), 1-4.

37. Porto, N. F., Fernández, J. D., Pallero, M. D. L. Á. G., Cuesta, J. R. P., Rivas, P. P., & Simoes, R. G. (2019). Metastasis in subcutaneous cellular tissue of glioblastoma multiforme: clinical case and literature review. *Neurocirugía*, *30*(3), 149-154.

38. Prelaj, A., Rebuzzi, S. E., Caffarena, G., Berrìos, G., Rodrigo, J., Pecorari, S., ... & Magliocca, F. M. (2018). Therapeutic approach in glioblastoma multiforme with primitive neuroectodermal tumor components: Case report and review of the literature. *Oncology letters*, *15*(5), 6641-6647.

39. Rajagopalan V, El Kamar FG, Thayaparan R, Grossbard ML. Bone marrow metastases from glioblastoma multiforme--A case report and review of the literature. *J Neurooncol*. 2005;72(2):157-161.

40. Ranjan, S., Quezado, M., Garren, N., Boris, L., Siegel, C., Neto, O. L. A., ... & Gilbert, M. R. (2018). Clinical decision making in the era of immunotherapy for high grade-glioma: report of four cases. *BMC cancer*, *18*(1), 239.

41. Richard, S. A., Ye, Y., Li, H., Ma, L., & You, C. (2018). Glioblastoma multiforme subterfuge as acute cerebral hemorrhage: A case report and literature review. *Neurology international*, *10*(1).

42. Romo, C. G., Palsgrove, D. N., Sivakumar, A., Elledge, C. R., Kleinberg, L. R., Chaichana, K. L., ... & Holdhoff, M. (2019). Widely metastatic IDH1-mutant glioblastoma with oligodendroglial features and atypical molecular findings: a case report and review of current challenges in molecular diagnostics. *Diagnostic pathology*, *14*(1), 16.

43. Rosen, J., Blau, T., Grau, S. J., Barbe, M. T., Fink, G. R., & Galldiks, N. (2018). Extracranial metastases of a cerebral glioblastoma: a case report and review of the literature. *Case reports in oncology*, *11*(2), 591-600.

44. Roviello, G., Petrioli, R., Cerase, A., Marsili, S., Miracco, C., Rubino, G. y Tini, P. (2013). A husband and wife with simultaneous presentation of glioblastoma multiforme: a case report. *Case reports in oncology*, *6* (3), 538-543.

45. Sajan, A., Hewitt, K., Soleiman, A., & Velayudhan, V. (2020). Pineal glioblastoma: Case report and literature review of an exceedingly rare etiology for pineal region mass. *Clinical Imaging*, *60*(1), 95-99.

46. Shen, CX, Wu, JF, Zhao, W., Cai, ZW, Cai, RZ y Chen, CM (2017). Primary spinal glioblastoma multiforme: a case report and literature review. *Medicine*, *96* (16).

47. Theeler BJ, Ellezam B, Yust-Katz S, Slopis JM, Loghin ME, de Groot JF. Prolonged survival in adult neurofibromatosis type I patients with recurrent high-grade gliomas treated with bevacizumab. *J Neurol*. 2014;261(8):1559-1564.

48. Thummalapalli R, Heumann T, Stein J, et al. Hemophagocytic Lymphohistiocytosis Secondary to PD-1 and IDO Inhibition in a Patient with Refractory Glioblastoma. *Case Rep Oncol*. 2020;13(2):508-514. Published 2020 May 12.

49. Tokuda, Y., Tamura, R., Ohara, K., Yoshida, K., & Sasaki, H. (2017). A case of glioblastoma resected immediately after administering bevacizumab: consideration on histopathological findings and safety of surgery. *Brain tumor pathology*, *34*(2), 98-102.

50. Uppar, A., Konar, S. K., Nandeesh, B. N., & Shukla, D. (2019). H3K27M-Positive Primary Spinal Glioblastoma Presenting with Hemorrhage—A Rare Clinical Entity. *World neurosurgery*, *126*, 223-227.

51. Van Seggelen, W. O., De Vos, F. Y., Röckmann, H., Van Dijk, M. R., & Verhoeff, J. J. (2019). Occurrence of an Abscopal Radiation Recall Phenomenon in a Glioblastoma Patient Treated with Nivolumab and Re-Irradiation. *Case Reports in Oncology*, *12*(3), 896-900.

52. Wang, L., Sun, J., Li, Z., Chen, L., Fu, Y., Zhao, L., ... & Lu, D. (2017). Gliosarcomas with the BRAF V600E mutation: a report of two cases and review of the literature. *Journal of clinical pathology*, *70*(12), 1079-1083.

53. Wang, Y., Song, S., Su, X., Wu, J., Dai, Z., Cui, D., ... & Wang, Z. (2018). Radiation-induced glioblastoma with rhabdoid characteristics following treatment for medulloblastoma: A case report and review of the literature. *Molecular and clinical oncology*, *9*(4), 415-418.

54. Watanabe N, Ishikawa E, Kohzuki H, et al. Malignant transformation of pleomorphic xanthoastrocytoma and differential diagnosis: case report. *BMC Neurol*. 2020;20(1):21. Published 2020 Jan 15.

55. Widjaja A, Mix H, Gölkel C, et al. Uncommon metastasis of a glioblastoma multiforme in liver and spleen. *Digestion*. 2000;61(3):219-222.

56. Woo, P. Y., Lam, T. C., Pu, J. K., Li, L. F., Leung, R. C., Ho, J. M., ... & Ng, H. K. (2019). Regression of BRAFV600E mutant adult glioblastoma after primary combined BRAF-MEK inhibitor targeted therapy: a report of two cases. *Oncotarget*, *10*(38), 3818.

57. Zhang, C., Yao, Y., Wang, Y., Chen, Z., Wu, J., Mao, Y. y Zhou, L. (2010). Temozolomide for adult brain stem glioblastoma: case report of a long-term survivor. *International Journal of Neuroscience* , *120* (12), 787-791.

58. Zhang, H., Chen, F., Wang, Z., & Wu, S. (2017). Successful treatment with apatinib for refractory recurrent malignant gliomas: a case series. *OncoTargets and therapy*, *10*, 837.

59. Zhou, K., Yao, H., Zhang, X., Liu, J., Qi, Z., Xie, X., ... & Che, Y. (2017). Next generation sequencing and molecular imaging identify EGFR mutation and amplification in a glioblastoma multiforme patient treated with an EGFR inhibitor: a case report. *Oncotarget*, *8*(30), 50305.

60. Zuccoli, G., Marcello, N., Pisanello, A., Servadei, F., Vaccaro, S., Mukherjee, P., y Seyfried, TN (2010). Metabolic management of glioblastoma multiforme using standard therapy together with a restricted ketogenic diet: case report. *Nutrition and metabolism*, *7* (1), 33.

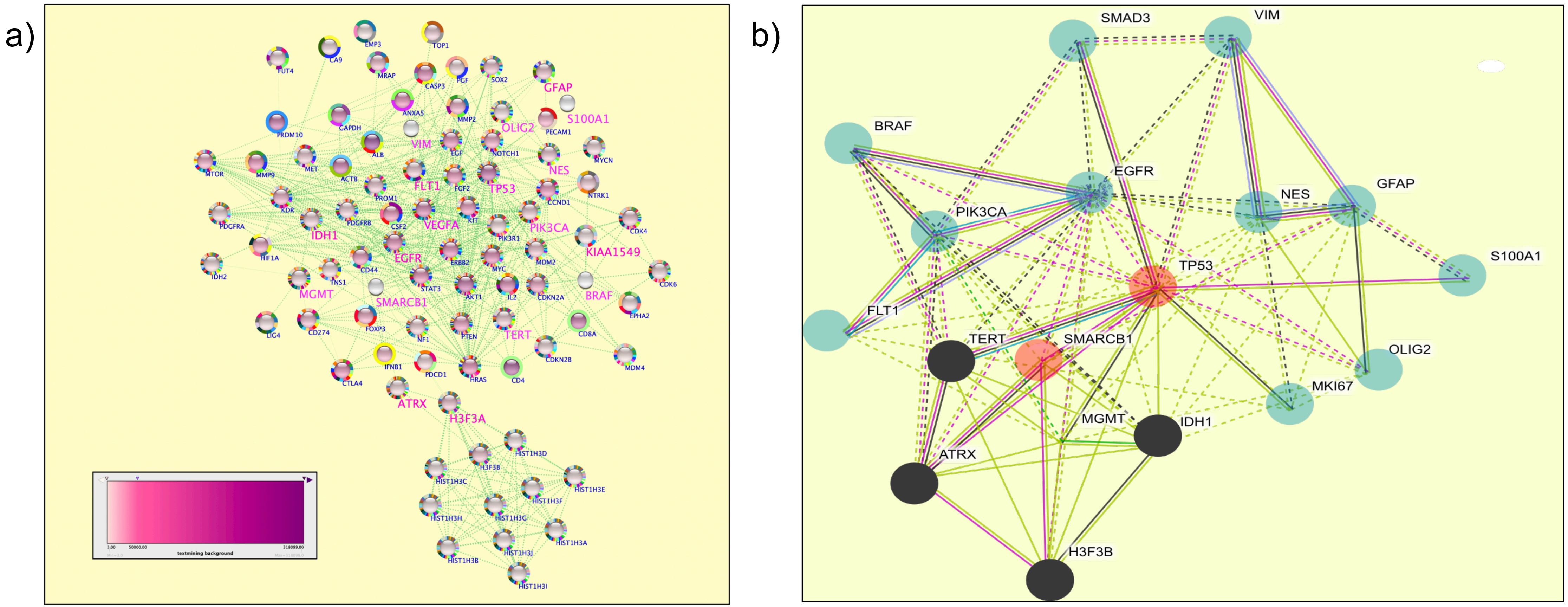


**Figure 2**. **Data mining selection and GO clustering of the most used genomic markers. a)** String network of the top genes reported in the PubMed Database, nodes are genes colored using predicted text mining score, references are indicated as spliced multicolor donuts, selected driver genes are highlighted in pink. B) GO clustering by K means of the GBM driver genes selected for meta-analysis. Nodes are genes colored by GO: BP groups, black: *DNA repair and chromatin remodeling,* red: *tumor suppressors,* blue: *cytoskeleton and cellular proliferation.* For both network edges represent protein-protein associations specific and meaningful, i.e. proteins jointly contribute to a shared function; this does not necessarily mean they are physically binding each other. Known Interactions from curated databases and experimentally determined are in green and pink, respectively.
